# miR487a-3p and miR6855-3p Facilitate Macrophage Pro-inflammatory Polarization and Lipid Accumulation in Atherosclerosis

**DOI:** 10.64898/2026.05.21.26353835

**Authors:** Haijing Ge, Duo Xu, Tao He, Zixing Zhang, Weina Wang, Jing Wan, Huanhuan Cai, Hairong Wang, Sumanth D. Prabhu, Zhibing Lu, Qiongxin Wang

## Abstract

**BACKGROUND:** The integrated regulation of microRNAs (miRNAs) on macrophage plasticity plays a key role in atherosclerosis (AS). We tested the hypothesis that miR487a-3p and miR6855-3p accelerate AS by intensifying macrophage inflammatory response and metabolic dysregulation.

**METHODS:** MiRNA sequencing (miR-seq) and mRNA sequencing (mRNA-seq) were conducted in peripheral monocytes from CAD patients and healthy controls. Macrophages in mouse aortas and human coronary arteries were characterized using flow cytometry and immunostaining. AS development was evaluated in male PCSK9-overexpression mice harboring myeloid cell-specific deficiency of carboxypeptidase E (CPE) or ribonucleotide reductase subunit M2 (RRM2) and challenged with a high-fat diet.

**RESULTS:** miR487a-3p and miR6855-3p were the top miRNA candidates identified by miR-seq in peripheral monocytes and validated by qPCR, with significant differences between coronary artery disease (CAD) patients and controls. Both miRNAs were lipid-inducible and secreted extracellularly. Krüppel-like factor 5 and interferon regulatory factor 1 bound to the promoter regions of miR-487a-3p and miR-6855-3p, respectively, to enhance their transcription. Accordingly, CAD patients exhibited significantly elevated plasma miR487a-3p and miR6855-3p levels compared with controls, which positively correlated with blood lipid levels and Gensini score (reflecting CAD severity and prognosis). The area under the receiver operating characteristic curve (≈0.83 for each) supported their diagnostic accuracy. Of note, miR487a-3p and miR6855-3p were predominantly expressed in coronary arterial macrophages. The dramatic expansion of miR-487a-3p⁺ and miR-6855-3p⁺ macrophages and the elevated expression of both miRNAs in coronary arteries were positively associated with lesion area in CAD patients. Mechanistically, transcriptomic analyses and functional assays revealed that elevated miR-487a-3p or miR-6855-3p promoted macrophage pro-inflammatory responses, lipid metabolic dysregulation, and foam cell formation. Conversely, inhibition of either miRNA alleviated ox-LDL-induced macrophage inflammatory responses and lipid metabolic dysfunction. Moreover, conditioned medium from miR487a-3p- or miR6855-3p-overexpressing macrophages promoted endothelial cell apoptosis, whereas this effect was attenuated when endothelial cells were exposed to medium from ox-LDL-treated macrophages with miRNA inhibition. Furthermore, integration of downregulated genes from monocyte and macrophage mRNA-seq with TargetScan-predicted targets identified CPE and RRM2 as targets of miR-487a-3p and miR-6855-3p, respectively. Direct binding was confirmed by dual-luciferase assays and miRNA pulldown. Overexpression of CPE or RRM2 partially reversed the detrimental effects of miR487a-3p and miR6855-3p, respectively, on macrophage phenotypic switching and metabolic dysregulation. Conversely, monocyte/macrophage-specific depletion of CPE or RRM2 aggravated AS progression in hypercholesterolemic mice by instigating macrophage inflammatory responses and lipid metabolic disturbance.

**CONCLUSIONS:** miR487a-3p and miR6855-3p fulfil the criteria of promising biomarkers for CAD diagnosis and prognosis. Mechanistically, they intensify inflammatory responses and disrupt lipid metabolism in macrophages, identifying both miRNAs as potential therapeutic targets for CAD.

## INTRODUCTION

MicroRNAs (miRNAs) are small non-coding RNA molecules present in all eukaryotic cells that regulate the expression of up to 60% of protein-coding genes by binding to complementary sequences on messenger RNAs (mRNAs), thereby suppressing mRNA translation and consequently reducing protein output. ^1^ Their ability to modulate mRNA stability and translational efficiency makes them important post-transcriptional regulators of diverse biological processes.^2^ miRNAs play essential roles in cardiovascular development and function, and a deregulated cardiac-enriched miRNA profile contributes critically to the pathogenesis of coronary artery disease (CAD)^3^ Recent studies have highlighted the potential of miRNAs as promising biomarkers and therapeutic targets for CAD,^4,5^ which results from atherosclerotic occlusion of the coronary arteries and remains the leading cause of death globally and a major contributor to disability^6^ Stable plasma levels of several miRNAs are strongly associated with CAD risk and are being explored as minimally invasive biomarkers for CAD diagnosis and prognosis; examples include miR21, miR3149, miR122-5p, miR370 and miR208a as diagnostic markers, and miR132, miR210, miR140-3p, miR423-5p and miR320a as prognostic markers.^7^ Moreover, modulation of a single miRNA by agomiR or antagomiR can alter the CAD disease process, for instance, antagomiR21, antagomiR33, miR30 mimic, and antagomiR143 significantly attenuate CAD-related cardiomyocyte death and improve cardiac function, underscoring their therapeutic potential in CAD.^3^

Atherosclerosis (AS) has a significant inflammatory component characterized by an imbalance between pro-inflammatory and regulatory signals.^8,9^ Monocytes and their descendent macrophages play central roles in disease initiation and progression.^10^ Triggered by inflammatory cytokines, monocytes transmigrate across the endothelium to the vascular intima, where they differentiate into macrophages and take up lipids, forming foam cells that constitute a major component of the atherosclerotic plaque.^10^ Plaque macrophages exhibit high heterogeneity and plasticity, which shape the evolving plaque microenvironment through excessive lipid accumulation, cytokine hyperactivation, hypoxia, apoptosis, and necroptosis. The metabolic and functional transitions of plaque macrophages in response to microenvironmental cues not only influence ongoing and subsequent inflammatory responses within lesions but also directly govern atherosclerotic progression or regression.^11,12^

Several miRNAs have been implicated in macrophage regulation during AS. miR124 overexpression may represent a promising therapeutic strategy for AS and CAD by inhibiting p38 and subsequent macrophage apoptosis.^13^ Conversely, miR6516-5p contributes to the loss of protective pericardial macrophages in CAD by suppressing CD36 and macrophage lipid uptake.^14^ Furthermore, inhibition of miR-33 and overexpression of miR-155 can prevent macrophage foam cell transformation and markedly reduce atherosclerotic plaque progression.^3,15^ Although these studies highlight the critical roles of individual miRNAs in macrophage functional regulation during CAD, a systemic understanding of miRNA-mediated modulation of the pleiotropic capacity of monocytes and macrophages, the central cell types in AS, remains lacking.

In this study, using miRNA sequencing (miR-seq) and histological analyses, we identified significantly elevated expression of miR-487a-3p and miR-6855-3p in plasma, peripheral monocytes, and coronary plaque macrophages of CAD patients, which positively correlated with blood lipid levels and CAD severity. Through macrophage transcriptome sequencing and functional assays, we elucidated the molecular mechanisms by which miR-487a-3p and miR-6855-3p drive macrophage phenotypic switching and metabolic reprogramming toward pro-inflammatory responses and excessive lipid retention, and identified CPE and RRM2 as their respective direct targets. Furthermore, using myeloid-specific Cpe- and Rrm2-deficient mice on a hypercholesterolemic background, we demonstrated that suppression of these targets accelerates AS in vivo, thereby establishing a mechanistic link between the miRNA-target gene axis and AS progression. These findings establish miR487a-3p and miR6855-3p as promising biomarkers for CAD diagnosis and prognosis and as potential therapeutic targets.

## MATERIALS AND METHODS

### Data Availability

All data supporting the findings of this study are included in the main text and Supplemental Material. All data, analytical methods, and materials are available from the corresponding author upon reasonable request. Detailed experimental materials and methods and a Major Resources Table are available in the Supplemental Material.

### Human Studies

After obtaining informed consent, venous blood was collected preoperatively from CAD patients and non-CAD controls undergoing coronary angiography. Coronary arterial tissue was procured from CAD patients undergoing coronary artery bypass graft (CABG) surgery. Healthy coronary arterial tissue was obtained from organ donors. All subjects and donors were from Zhongnan Hospital of Wuhan University; clinical characteristics were presented in Table 1, 2, 4, 5 and S1, S2, S4. All protocols for collecting and using human blood and coronary arterial samples were approved by the Medical Ethics Committee of Zhongnan Hospital of Wuhan University (protocol No. 2025058K). Blood samples were used for the isolation of plasma and immune cells, and coronary arterial samples were used for immune cell isolation and subsequent flow cytometry, as well as histological analysis.

**Table 1.**
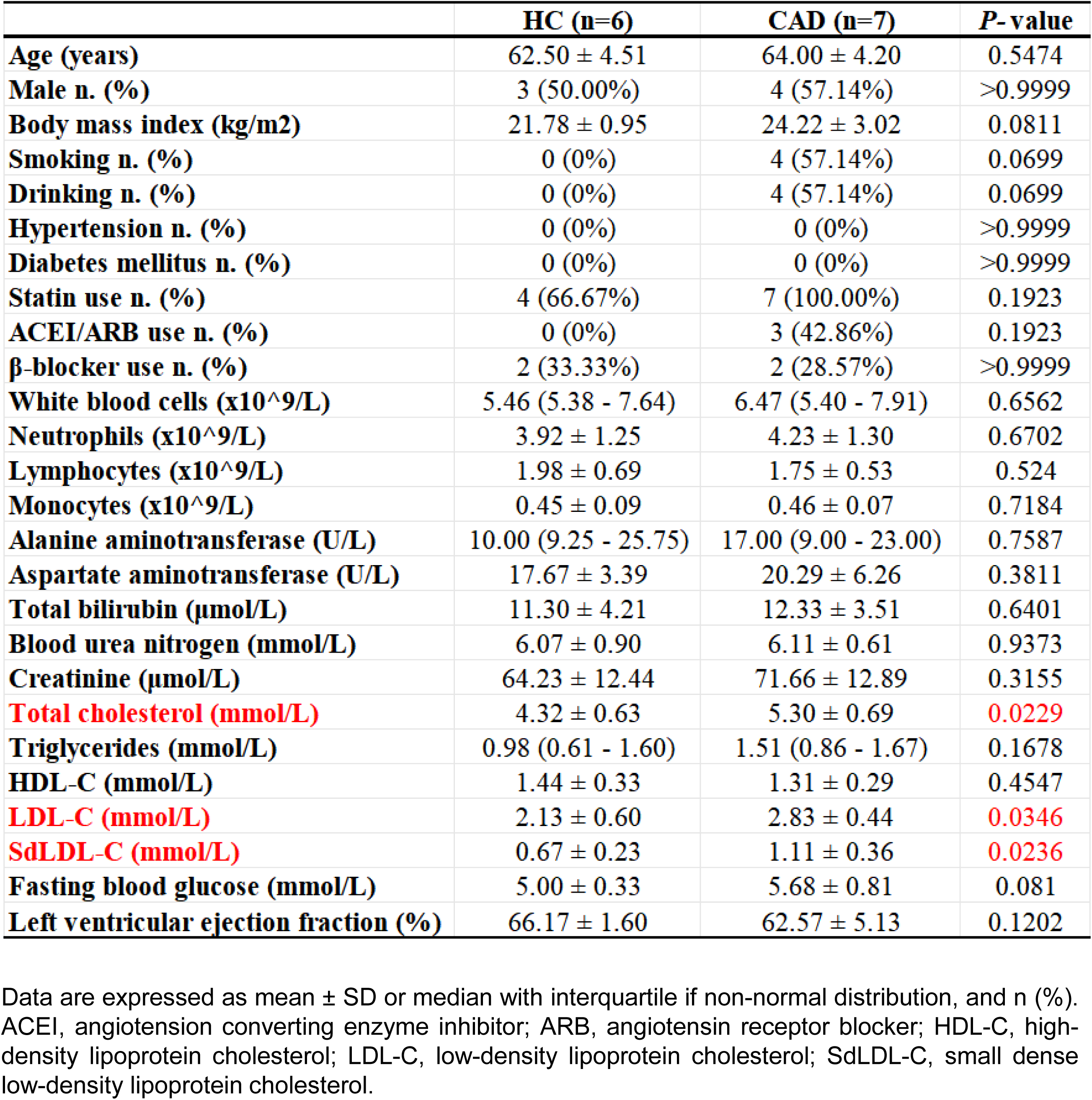
Demographic data of the health controls (HCs) and coronary artery disease (CAD) patients for monocyte miR-seq.

|  | **HC (n=6)** | **CAD (n=7)** | **P- value** |
| --- | --- | --- | --- |
| **Age (years)** | 62.50 ± 4.51 | 64.00 ± 4.20 | 0.5474 |
| **Male n. (%)** | 3 (50.00%) | 4 (57.14%) | >0.9999 |
| **Body mass index (kg/m2)** | 21.78 ± 0.95 | 24.22 ± 3.02 | 0.0811 |
| **Smoking n. (%)** | 0 (0%) | 4 (57.14%) | 0.0699 |
| **Drinking n. (%)** | 0 (0%) | 4 (57.14%) | 0.0699 |
| **Hypertension n. (%)** | 0 (0%) | 0 (0%) | >0.9999 |
| **Diabetes mellitus n. (%)** | 0 (0%) | 0 (0%) | >0.9999 |
| **Statin use n. (%)** | 4 (66.67%) | 7 (100.00%) | 0.1923 |
| **ACEI/ARB use n. (%)** | 0 (0%) | 3 (42.86%) | 0.1923 |
| **β-blocker use n. (%)** | 2 (33.33%) | 2 (28.57%) | >0.9999 |
| **White blood cells (x10<sup>9</sup>/L)** | 5.46 (5.38 - 7.64) | 6.47 (5.40 - 7.91) | 0.6562 |
| **Neutrophils (x10<sup>9</sup>/L)** | 3.92 ± 1.25 | 4.23 ± 1.30 | 0.6702 |
| **Lymphocytes (x10<sup>9</sup>/L)** | 1.98 ± 0.69 | 1.75 ± 0.53 | 0.524 |
| **Monocytes (x10<sup>9</sup>/L)** | 0.45 ± 0.09 | 0.46 ± 0.07 | 0.7184 |
| **Alanine aminotransferase (U/L)** | 10.00 (9.25 - 25.75) | 17.00 (9.00 - 23.00) | 0.7587 |
| **Aspartate aminotransferase (U/L)** | 17.67 ± 3.39 | 20.29 ± 6.26 | 0.3811 |
| **Total bilirubin (μmol/L)** | 11.30 ± 4.21 | 12.33 ± 3.51 | 0.6401 |
| **Blood urea nitrogen (mmol/L)** | 6.07 ± 0.90 | 6.11 ± 0.61 | 0.9373 |
| **Creatinine (μmol/L)** | 64.23 ± 12.44 | 71.66 ± 12.89 | 0.3155 |
| **Total cholesterol (mmol/L)** | 4.32 ± 0.63 | 5.30 ± 0.69 | **0.0229** |
| **Triglycerides (mmol/L)** | 0.98 (0.61 - 1.60) | 1.51 (0.86 - 1.67) | 0.1678 |
| **HDL-C (mmol/L)** | 1.44 ± 0.33 | 1.31 ± 0.29 | 0.4547 |
| **LDL-C (mmol/L)** | 2.13 ± 0.60 | 2.83 ± 0.44 | **0.0346** |
| **SdLDL-C (mmol/L)** | 0.67 ± 0.23 | 1.11 ± 0.36 | **0.0236** |
| **Fasting blood glucose (mmol/L)** | 5.00 ± 0.33 | 5.68 ± 0.81 | 0.081 |
| **Left ventricular ejection fraction (%)** | 66.17 ± 1.60 | 62.57 ± 5.13 | 0.1202 |

Data are expressed as mean ± SD or median with interquartile if non-normal distribution, and n (%). ACEI, angiotension converting enzyme inhibitor; ARB, angiotensin receptor blocker; HDL-C, high-density lipoprotein cholesterol; LDL-C, low-density lipoprotein cholesterol; SdLDL-C, small dense low-density lipoprotein cholesterol.

### Animal Studies

All animal experiments complied with the National Institutes of Health Guide for the Care and Use of Laboratory Animals, and were performed under institutional Animal Care and Use Committee protocol no. ZN2023042 at Zhongnan Hospital of Wuhan University. Studies conformed to the ARRIVE (Animal Research: Reporting of In Vivo Experiments) 2.0 reporting guidelines. Male mice at 7-w old were used for experiments as indicated.

### Data Analysis

Data are expressed as mean±SD, or median with interquartile if non-normal distribution. Normality was assessed using the Shapiro-Wilk test, and group variances were compared using F test for datasets with 2 groups, or the Brown-Forsythe test with groups of 3 or more. For normally distributed data, unpaired Student’s t test with equal or unequal variance (Welch’s correction) was used for two-group analyses, and 1- or 2-way ANOVA test was used for multi-group analyses. For non-normal distributions, the Mann-Whitney *U* test was used for 2-group analyses. Categorical data were analyzed using the Fisher’s exact test and presented as counts (percentages). Pearson or Spearman (for non-normal distributions) correlation analysis was used to assess the correlation between the 2 indicators. Univariate and multivariate logistic regression analyses were performed to evaluate the independent association of plasma miRNA levels and lipid parameters with CAD status. Specific approaches are detailed in the figure legends. All statistical analyses were performed with GraphPad Prism 10.4.0 (GraphPad Software). A p value of < 0.05 was considered statistically significant.

## RESULTS

### miR487a-3p and miR6855-3p Are Upregulated in Monocytes and Plasma of CAD Patients and Correlate with Atherosclerosis Severity

To identify miRNAs dysregulated in CAD, we performed miR-seq on peripheral monocytes from CAD patients and healthy controls (HCs) (Figure 1A). The purity of negatively selected monocytes from PBMCs of non-hypertensive and non-diabetic subjects reached ∼90% (Figure 1B). Blood lipid levels, including total cholesterol (TC), low-density lipoprotein cholesterol (LDL-C), and small dense LDL-C (sdLDL-C), were significantly higher in CAD patients compared with HCs, whereas other biochemical parameters, hematimetry, and left ventricular ejection fraction were comparable between groups (Table 1). A total of 50 miRNAs were differentially expressed in monocytes from CAD patients versus HCs (Figure 1C). A heatmap of the top 17 miRNAs (|log2FC| ≥ 1.9) revealed 6 markedly upregulated and 11 downregulated miRNAs (Figure 1D). GO and KEGG enrichment analyses of the target genes of these 50 miRNAs revealed pathways related to inflammation, immune activation, and lipid metabolism (Figure S1A and S1B).

**Figure 1.**
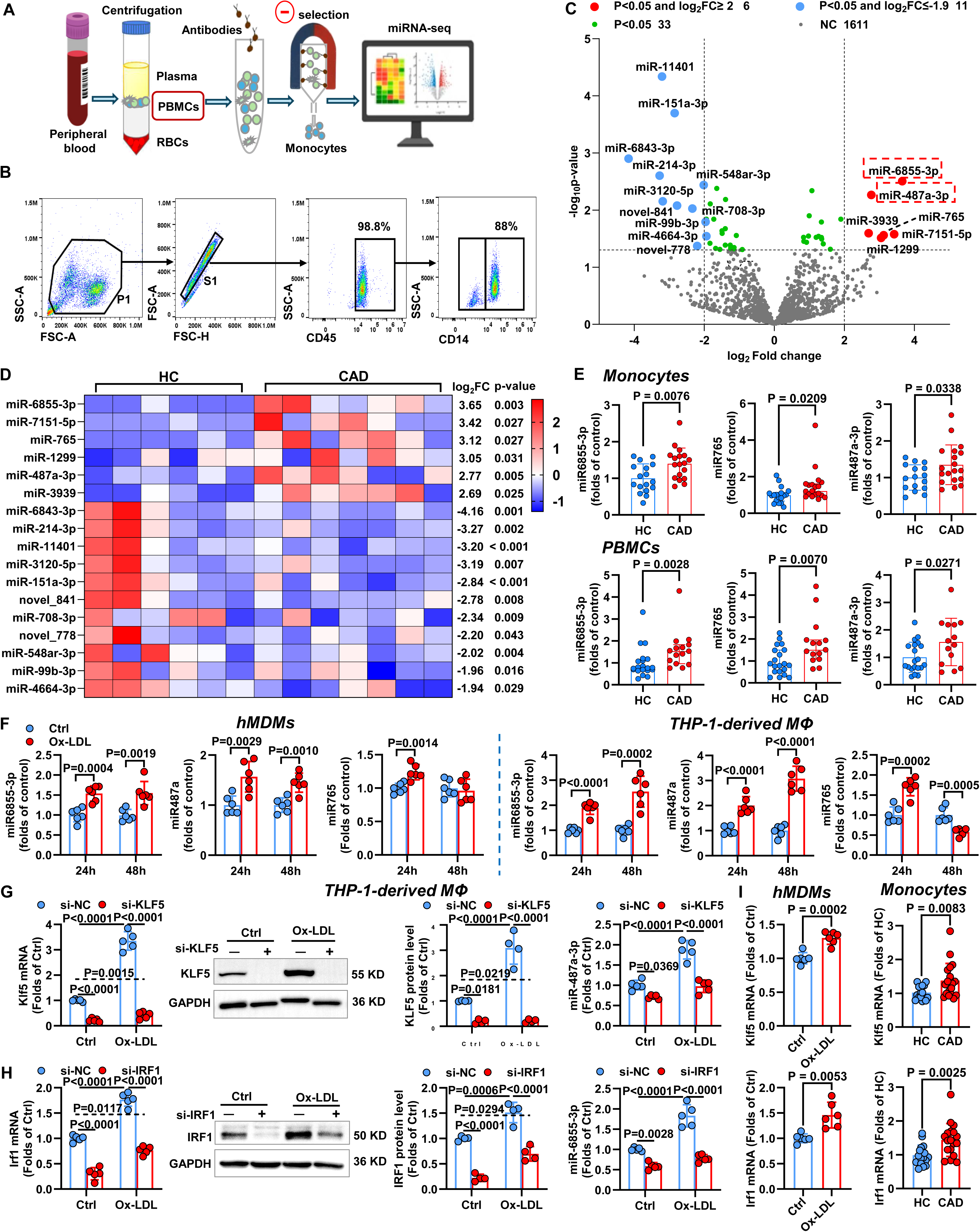
miR487a-3p and miR6855-3p are increased in peripheral monocytes of patients with coronary artery disease (CAD) compared to healthy controls (HCs). **A**, Schematic showing the process of peripheral blood mononuclear cells (PBMCs) collection, monocyte isolation, and microRNA-sequencing (miR-seq) on Illumina platforms. **B**, Representative flow cytometry plots showing peripheral leukocytes (CD45^+^) and monocytes (CD14^+^). **C**, Volcano plot displaying differentially expressed miRNAs in peripheral monocytes from HCs (n=6) and CAD patients (n=7). **D**, Heatmap showing expression levels of the 17 differentially expressed miRNAs (red and blue dots in *C*). **E**, The expressions of miR6855-3p, miR487a-3p and miR765 in peripheral monocytes (HC: 18, CAD: 19) and PBMCs (HC: 20, CAD: 15) from HCs and CAD patients. **F**, The expressions of miR6855-3p, miR487a-3p and miR765 in human monocyte-derived macrophages (hMDMs) (Donor: 6) and THP-1-derived macrophages treated with vehicle (Ctrl) or 100 μg/mL oxidized low-density lipoprotein (ox-LDL) for 24 or 48 h (n=6). **G-H**, The mRNA and protein levels of KLF5 (*G*) or IRF1 (*H*) and the expressions of miR487a-3p (*G*) or miR6855-3p (*H*) in THP-1-derived macrophages transfected with negative control (NC) or KLF5 (*G*) or IRF1 (*H*) siRNA, followed by treatment with Ctrl or 100 μg/mL ox-LDL for 24 h. **I**, mRNA expressions of Klf5 and Irf1 in hMDMs (Donor: 6) treated with Ctrl or ox-LDL for 24 h or in peripheral monocytes from HCs (n=17) and CAD patients (n=19). Statistics: Unpaired Student’s *t*-test in *E*, *F* and *I*, and Mann-Whitney *U* test in *E* and *I*. 2-way ANOVA with post-hoc Tukey’s multiple comparisons between groups in *G* and *H*.

To validate these findings, we measured the expression of the top 17 miRNAs in an expanded cohort of peripheral monocytes and PBMCs from HCs and CAD patients (Tables S1 and S2). Among these, miR6855-3p, miR765, and miR487a-3p were consistently upregulated in both PBMCs and monocytes from CAD patients compared with HCs, confirming the miR-seq results (Figure 1E; S2A and S2B).

Given the hyperlipidemic profile of CAD patients, we next tested whether these three miRNAs were responsive to lipid stimulation. In both human monocyte-derived macrophages (hMDMs) and THP-1-derived macrophages, ox-LDL robustly induced the expression of miR487a-3p and miR6855-3p, but not miR765, over 48 h (Figure 1F). This lipid-inducible expression pattern led us to focus on miR487a-3p and miR6855-3p for further investigation, excluding miR765. To identify the upstream regulators mediating this induction, we used the TransmiR database to screen candidate transcription factors (TFs). Among 35 candidate TFs for miR487a-3p, only two were predicted to activate its transcription (Figure S3A), and ChIP assay confirmed the binding of Krüppel-like factor 5 (KLF5) to the miR487a-3p promoter (Figure S3C). Ox-LDL markedly induced KLF5 expression in THP-1-derived macrophages, and KLF5 silencing significantly reduced miR487a-3p expression both at baseline and under ox-LDL stimulation (Figure 1G; S3B), establishing KLF5 as the mediator of ox-LDL-induced miR487a-3p transcription. For miR6855-3p, parallel bioinformatic screening and gene expression validation identified interferon regulatory factor 1 (IRF1) and early growth response 1 (EGR1) as candidate TFs (Figure S3D), both of which were activated by ox-LDL (Figure 1H; S3E and S3F). Silencing of IRF1, but not EGR1, significantly reduced miR6855-3p expression under both basal and ox-LDL-stimulated conditions (Figure 1H; S3F), identifying IRF1 as the primary TF driving ox-LDL-induced miR6855-3p transcription, with the predicted binding site shown in Figure S3H.

Given that cellular miRNAs can be released into the extracellular environment and are detectable in biofluids including plasma, follicular fluid and even in cell culture media.^16,17^ we tested whether miR487a-3p and miR6855-3p were secreted from macrophages. Both miRNAs were robustly released into the supernatant of THP-1-derived macrophages and hMDMs upon ox-LDL treatment, paralleling their intracellular upregulation (Figure S4A and S4B). Moreover, silencing of KLF5 or IRF1 significantly reduced their extracellular release (Figure S4A), indicating that intracellular upregulation in macrophages contributes to their extracellular levels. We therefore measured plasma miR487a-3p and miR6855-3p levels in a large cohort of CAD patients and controls who had comparable demographic characteristics but differed in blood lipid levels and medication history (Table 2, more anti-hypertensive and cholesterol-lowering drug use in CAD vs. control). Plasma levels of both miRNAs were significantly elevated in CAD patients compared with controls and positively correlated with blood TC, LDL-C, and sdLDL-C, but not with triglycerides, high-sensitivity C-reactive protein, blood pressure, or fasting blood glucose (Figure 2A; S4C and S4D). Both miRNAs were comparable between smoking and non-smoking CAD patients (Figure S4E). Given that sdLDL-C is the most atherogenic lipoprotein parameter and both sdLDL-C and LDL-C contribute significantly to atherosclerotic cardiovascular disease,^18,19^ these results suggest a link between elevated circulating miR487a-3p and miR6855-3p and AS in CAD patients.

**Figure 2.**
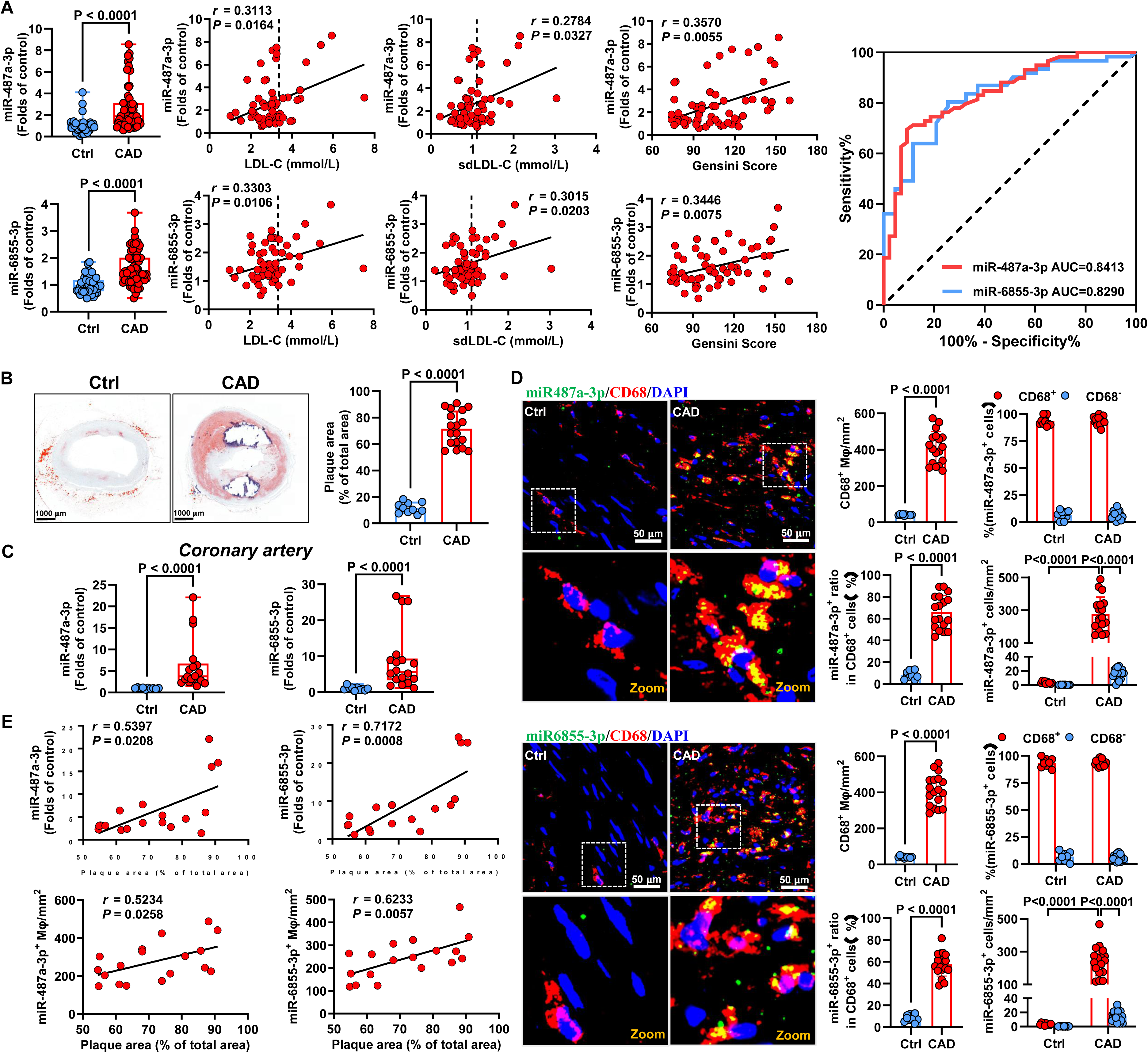
Increases of miR487a-3p and miR6855-3p in plasma and coronary arteries of patients with coronary artery disease (CAD) correlate with hyperlipidemia and plaque area respectively. **A**, Plasma miR487a-3p and miR6855-3p levels in controls (Ctrls) and CAD patients (left). Correlations between plasma miR487a-3p or miR6855-3p and low-density lipoprotein cholesterol (LDL-C), small dense LDL-C (sdLDL-C), and Gensini score in CAD patients (middle). Receiver operating characteristic (ROC) curves for evaluating the diagnostic value of miR487a-3p and miR6855-3p for CAD (right). Ctrl, n=43; CAD, n=59. **B**, Representative Oil Red O staining of coronary artery sections from Ctrls and CAD patients, with quantitation of plaque area ratios. Scale bar 100 μm. **C**, The expressions of miR487a-3p and miR6855-3p in coronary arteries of Ctrls and CAD patients. **D**, Representative low- and high-magnification confocal images of coronary arteries from Ctrls and CAD patients stained for CD68 (red) and fluorescent miR487a-3p and miR6855-3p in situ hybridization (green), with quantitation for the numbers and frequencies of CD68^+^, miR487a-3p^+^ and miR6855-3p^+^ cells. DAPI (blue) was used to label nuclei. Scale bar 50 μm. Ctrls (n=10) and CAD (n=18) in *B*-*D*. **E**, Correlations between plaque area ratios and coronary artery miR487a-3p or miR6855-3p levels, and miR487a-3p^+^ or miR6855-3p^+^ macrophage abundance in CAD patients (n=18). Statistics: Unpaired Student’s *t*-test in *D*, Welch’s *t*-test in *B* and *D*, and Mann-Whitney *U* test in *A*, *C*, and *D*. Pearson correlation analysis in *E*, and Spearman correlation analysis in *A* and *E*.

**Table 2.** Demographic data of the controls (Ctrls) and coronary artery disease (CAD) patients for plasma miRNA expression assay.

|  | **Ctrl (n=43)** | **CAD (n=59)** | **P- value** |
| --- | --- | --- | --- |
| **Age (years)** | 61.12 ± 6.38 | 63.73 ± 9.13 | 0.0921 |
| **Male n. (%)** | 25 (58.14%) | 44 (74.58%) | 0.0905 |
| **Body mass index (kg/m2)** | 23.44 (21.48 - 25.39) | 24.80 (22.72 - 26.22) | 0.1499 |
| **Smoking n. (%)** | 9 (20.93%) | 18 (30.51%) | 0.3646 |
| **Drinking n. (%)** | 5 (11.63%) | 13 (22.03%) | 0.199 |
| **Hypertension n. (%)** | 7 (16.28%) | 19 (32.20%) | 0.1063 |
| **Diabetes mellitus n. (%)** | 3 (6.98%) | 11 (18.64%) | 0.144 |
| **Statin use n. (%)** | 36 (83.72%) | 59 (100%) | **0.0017** |
| **ACEI/ARB use n. (%)** | 2 (4.65%) | 29 (49.15%) | **<0.0001** |
| **β-blocker use n. (%)** | 5 (11.63%) | 30 (50.85%) | **<0.0001** |
| **White blood cells (x10<sup>9</sup>/L)** | 5.91 (5.30 - 6.60) | 6.07 (5.10 - 7.20) | 0.6597 |
| **Neutrophils (x10<sup>9</sup>/L)** | 3.61 (3.20 - 4.12) | 3.80 (3.20 - 4.80) | 0.4773 |
| **Lymphocytes (x10<sup>9</sup>/L)** | 1.67 ± 0.51 | 1.55 ± 0.44 | 0.2059 |
| **Monocytes (x10<sup>9</sup>/L)** | 0.45 (0.40 - 0.51) | 0.50 (0.40 - 0.60) | 0.1558 |
| **Alanine aminotransferase (U/L)** | 17.00 (11.00 - 27.00) | 19.00 (14.00 - 26.00) | 0.2767 |
| **Aspartate aminotransferase (U/L)** | 18.00 (16.00 - 23.00) | 19.00 (17.00 - 24.00) | 0.3269 |
| **Total bilirubin (μmol/L)** | 12.80 (10.00 - 16.20) | 11.70 (8.30 - 14.30) | 0.1712 |
| **Blood urea nitrogen (mmol/L)** | 5.50 (4.80 - 7.07) | 6.40 (5.20 - 7.50) | 0.1096 |
| **Creatinine (μmol/L)** | 67.80 (58.50 - 86.30) | 73.60 (66.00 - 84.90) | 0.1578 |
| **Total cholesterol (mmol/L)** | 4.12 (3.53 - 4.75) | 5.20 (4.20 - 5.42) | **<0.0001** |
| **Triglycerides (mmol/L)** | 1.14 (0.90 - 1.55) | 1.40 (1.13 - 1.61) | 0.0658 |
| **HDL-C (mmol/L)** | 1.09 (0.96 - 1.35) | 1.09 (0.93 - 1.24) | 0.6402 |
| **LDL-C (mmol/L)** | 2.36 (1.85 - 2.77) | 3.01 (2.53 - 3.37) | **<0.0001** |
| **SdLDL-C (mmol/L)** | 0.73 (0.49 - 0.96) | 1.00 (0.79 - 1.23) | **0.0004** |
| **Fasting blood glucose (mmol/L)** | 5.32 (5.00 - 5.63) | 5.52 (4.90 - 6.43) | 0.2803 |
| **Left ventricular ejection fraction (%)** | 65.00 (62.00 - 67.00) | 64.00 (60.00 - 68.00) | 0.3943 |

Data are expressed as mean ± SD or median with interquartile if non-normal distribution, and n (%). ACEI, angiotension converting enzyme inhibitor; ARB, angiotensin receptor blocker; HDL-C, high-density lipoprotein cholesterol; LDL-C, low-density lipoprotein cholesterol; SdLDL-C, small dense low-density lipoprotein cholesterol.

To further evaluate the clinical relevance of these circulating miRNAs, we examined their relationship with coronary AS severity using the Gensini score, a quantitative measure of total coronary atherosclerotic burden that provides valuable information on CAD severity and prognosis.^20,21^ Plasma miR487a-3p and miR6855-3p levels positively correlated with the Gensini score (Figure 2A). ROC analysis was performed to assess their diagnostic performance. The area under the ROC curve (AUC) for miR487a-3p was 0.841, with a sensitivity of 76.27% and specificity of 76.74% (cut-off value, 1.175), while the AUC for miR6855-3p was 0.829, with a sensitivity of 80.33% and specificity of 74.42% (cut-off value, 1.155) (Figure 2A). These results demonstrate the reliability of circulating miR-487a-3p and miR-6855-3p as diagnostic biomarkers for CAD. To assess whether miR487a-3p and miR6855-3p are independent predictors of CAD beyond their association with lipid profiles, we performed single-factor and multi-factor logistic regression analyses. In univariate analysis, miR487a-3p (OR=4.322, P=0.0001), miR6855-3p (OR=22.258, P<0.0001), TC (OR=2.89, P=0.0001), LDL-C (OR=3.417, P=0.0003), and sdLDL-C (OR=4.935, P=0.0061) were all significantly associated with CAD (Table S3). In multivariate logistic regression incorporating all three lipid parameters, both miRNAs remained significant independent predictors: miR487a-3p (OR=2.356, P=0.0202) and miR6855-3p (OR= 6.718, P=0.0161), whereas none of the lipid parameters retained significance (Table 3). These data demonstrate that plasma miR487a-3p and miR6855-3p are independent risk factors for CAD and that their predictive value is not merely a reflection of circulating lipid levels.

**Table 3.** Multivariate logistic regression analysis of plasma miRNA levels and lipid parameters associated with coronary artery disease.

| Variables | B | S.E. | Wald | *P*-value | OR | 95% CI |
| --- | --- | --- | --- | --- | --- | --- |
| miR-487a-3p | 0.86 | 0.37 | 5.40 | 0.0202 | 2.36 | 1.14 - 4.86 |
| miR-6855-3p | 1.90 | 0.79 | 5.80 | 0.0161 | 6.72 | 1.43 - 31.67 |
| TC | 0.28 | 0.51 | 0.31 | 0.5772 | 1.33 | 0.49 - 3.57 |
| LDL-C | 0.60 | 0.65 | 0.87 | 0.3511 | 1.82 | 0.52 - 6.46 |
| sdLDL-C | 0.26 | 0.83 | 0.09 | 0.7587 | 1.29 | 0.25 - 6.59 |

B, unstandardized coefficient; S.E. , standard error; Wald, Wald chi-square statistic; OR, odds ratio; CI, confidence interval; LDL-C, low-density lipoprotein cholesterol; sdLDL-C, small dense low-density lipoprotein cholesterol.

### miR487a-3p and miR6855-3p Are Enriched in Coronary Plaque Macrophages and Correlate with Lesion Area

To further understand the translational relevance of these findings, we examined the expression of miR487a-3p and miR6855-3p in coronary atherosclerotic plaques, where macrophages derived from infiltrating monocytes play a central role.^10,11,22,23^ Coronary arterial tissue was procured from patients with severe CAD undergoing CABG and from control donor hearts unsuitable for transplantation (Table 4). Oil Red O staining revealed that plaque area occupied approximately 72% of the coronary arterial wall in the CAD group, compared with 11% in controls (Figure 2B). Accordingly, miR487a-3p and miR6855-3p expression was dramatically increased in the coronary arteries of CAD patients versus controls (Figure 2C). Consistent with previous reports,^8,11,23^ macrophages (CD68^+^) were significantly expanded (∼10-fold) in the arterial wall of CAD patients compared with controls (Figure 2D; S4F).

**Table 4.** Demographic data of the controls (Ctrls) and coronary artery disease (CAD) patients for miRNA expression assay in coronary arteries.

|  | **Ctrl (n=10)** | **CAD (n=18)** | ***P*-value** |
| --- | --- | --- | --- |
| **Age (years)** | 57.60 ± 5.54 | 58.39 ± 7.97 | 0.7840 |
| **Male n. (%)** | 8 (80.00%) | 14 (77.78%) | >0.9999 |
| **Body mass index (kg/m<sup>2</sup>)** | 23.74 ± 2.02 | 22.64 ± 2.44 | 0.2379 |
| **Smoking n. (%)** | 0 (0%) | 1 (5.56%) | >0.9999 |
| **Drinking n. (%)** | 0 (0%) | 0 (0%) | >0.9999 |
| **Hypertension n. (%)** | 2 (20.00%) | 8 (44.44%) | 0.2474 |
| **Diabetes mellitus n. (%)** | 0 (0%) | 3 (16.67%) | 0.5330 |
| **Statin use n. (%)** | 0 (0%) | 18 (100%) | **<0.0001** |
| **ACEI/ARB use n. (%)** | 0 (0%) | 1 (5.56%) | >0.9999 |
| **β-blocker use n. (%)** | 1 (10.00%) | 7(38.89%) | 0.1937 |
| **Total cholesterol (mmol/L)** | — | 5.42 ± 0.37 | — |
| **Triglycerides (mmol/L)** | — | 1.60 ± 0.51 | — |
| **HDL-C (mmol/L)** | — | 1.12 ± 0.28 | — |
| **LDL-C (mmol/L)** | — | 3.43 ± 0.42 | — |
| **SdLDL-C (mmol/L)** | — | 1.12 ± 0.24 | — |

Data are expressed as mean ± SD and n (%). ACEI, angiotension converting enzyme inhibitor; ARB, angiotensin receptor blocker; HDL-C, high-density lipoprotein cholesterol; LDL-C, low-density lipoprotein cholesterol; SdLDL-C, small dense low-density lipoprotein cholesterol. For organ donor ctrls, part of clinical data were not available.

We next determined the cellular source of these miRNAs using fluorescence in situ hybridization (FISH) combined with immunofluorescence staining for CD68. Macrophages constituted approximately 93% of coronary arterial miR487a-3p^+^ or miR6855-3p^+^ cells in both groups. However, the proportion of macrophages expressing these miRNAs differed markedly: in CAD patients, 66.1% of macrophages were miR487a-3p^+^ and 57.7% were miR6855-3p^+^, compared with only 8.3% and 8.4% in controls, respectively (Figure 2D). Consequently, miR487a-3p^+^ and miR6855-3p^+^ macrophages were massively expanded (>30-fold vs. controls) in the coronary arterial wall of CAD patients, accounting for the elevated expression of both miRNAs in CAD coronary tissue. In contrast, miR487a-3p^+^ and miR6855-3p^+^ macrophages were barely detectable in control arteries, where few macrophages were present and both miRNAs were expressed at low levels (Figure 2D; S4F).

Notably, coronary plaque area in CAD patients was positively correlated with both arterial miR487a-3p and miR6855-3p expression and with the abundance of miR487a-3p^+^ and miR6855-3p^+^ macrophages in the arterial wall (Figure 2E). Together, these results demonstrate that miR487a-3p and miR6855-3p are predominantly expressed in arterial macrophages, that miR-487a3p^+^ and miR6855-3p^+^ macrophages expand robustly in atherosclerotic plaques, and that their abundance is associated with lesion burden, implicating macrophage-derived miR487a-3p and miR6855-3p in atherogenesis.

### miR-487a-3p and miR-6855-3p Reprogram Macrophages Toward Pro-Inflammatory and Pro-Atherogenic Gene Expression

To investigate how miR487a-3p and miR6855-3p modulate macrophage function in atherogenesis, we performed mRNA sequencing (mRNA-seq) on THP-1-derived macrophages transfected with miR487a-3p mimic, miR6855-3p mimic or negative control (NC) for 24 h (Figure S5A and S6A).

miR-487a-3p overexpression induced 2,852 gene upregulation and 3,112 gene downregulation versus NC (Figure 3A). Gene Set Enrichment Analysis (GSEA)-based GO analysis revealed enrichment of terms related to pro-inflammatory responses, including positive regulation of the mitogen-activated protein kinase (MAPK) cascade, Nuclear Factor Kappa B (NF-κB) signaling, cytokine release and binding, chemotaxis, and immune cell activation, as well as pro-atherogenic processes such as apoptosis, response to lipid, and suppression of lipid storage and transport and extracellular matrix organization (Figure 3B). GSEA-based KEGG analysis further demonstrated upregulation of pathways involved in lipid metabolism and atherosclerosis, chemokine and cytokine–cytokine receptor interactions, and inflammatory signaling pathways including Toll-like receptor, tumor necrosis factor (TNF), NOD-like receptor, Janus kinase/signal transduction and transcription activation (JAK/STAT), NF-κB, and MAPK (Figure 3C). Heatmaps displaying DEGs by functional category (Figure 3D and 3E; S5B and S5C) were validated by RT-qPCR (quantitative reverse transcription polymerase chain reaction) (Figure S5D–S5F). C-C and C-X-C motif chemokine ligands (CCLs and CXCLs) and their receptors (CCRs and CXCRs), orchestrate monocyte and macrophage mobilization, recruitment, and fate during AS initiation and progression.^24–26^ miR-487a-3p overexpression upregulated cytokines such as colony-stimulating factor (Csf) 1, Csf2, Csf3, interleukin (IL) 1a, Il15, Il27, and TNF superfamily (TNFSF) 10, and chemokines (Ccl3, Ccl3l1, Ccl4, Ccl5, Ccl8, Ccl20 and Cxcl1, Cxcl3, Cxcl8, Cxcl9, Cxcl10, Cxcl11, Cxcl12, Cxcl13) and their receptors (Ccr1, Ccr4, Ccr7, Ccrl2 and Cxcr4, Cxcr5) (Figure 3D; S5D). Pro-inflammatory polarization markers, including Il6, Ccl4l2, Cd80, Tnf, Il1b, interferon (IFN) b1 and Ccl2), were upregulated, whereas anti-inflammatory markers, such as CD206 (Mrc1), Cd163, endothelial growth factor (VEGF) b and Il4r, were downregulated (Figure 3E; S5E). Furthermore, cholesterol efflux genes (Apoe, Abca1, Abcg1, Scarb1 and Scarb2) were suppressed while the cholesterol uptake receptor Olr1 was increased, with only a mild decrease in Cd36 (Figure 3E; S5E), indicating a net shift toward lipid accumulation. Additionally, miR487a-3p perturbed extracellular matrix homeostasis by downregulating tissue inhibitor of matrix metalloproteinase (Timp) 1, Timp2, Timp3 and Mmp2, Mmp7, Mmp9, Mmp14, Mmp17, and upregulating matrix metalloproteinase (Mmp) 3, Mmp8, Mmp10, Mmp13 (Figure S5B). Key transcription factors were also altered, with upregulation of Klf2, Klf4, Klf5, and Nfkb1, and downregulation of Srebf1, Srebf2, Ppara, Pparg, Rhoa, and Rxra (Figure S5C and S5F).

**Figure 3.**
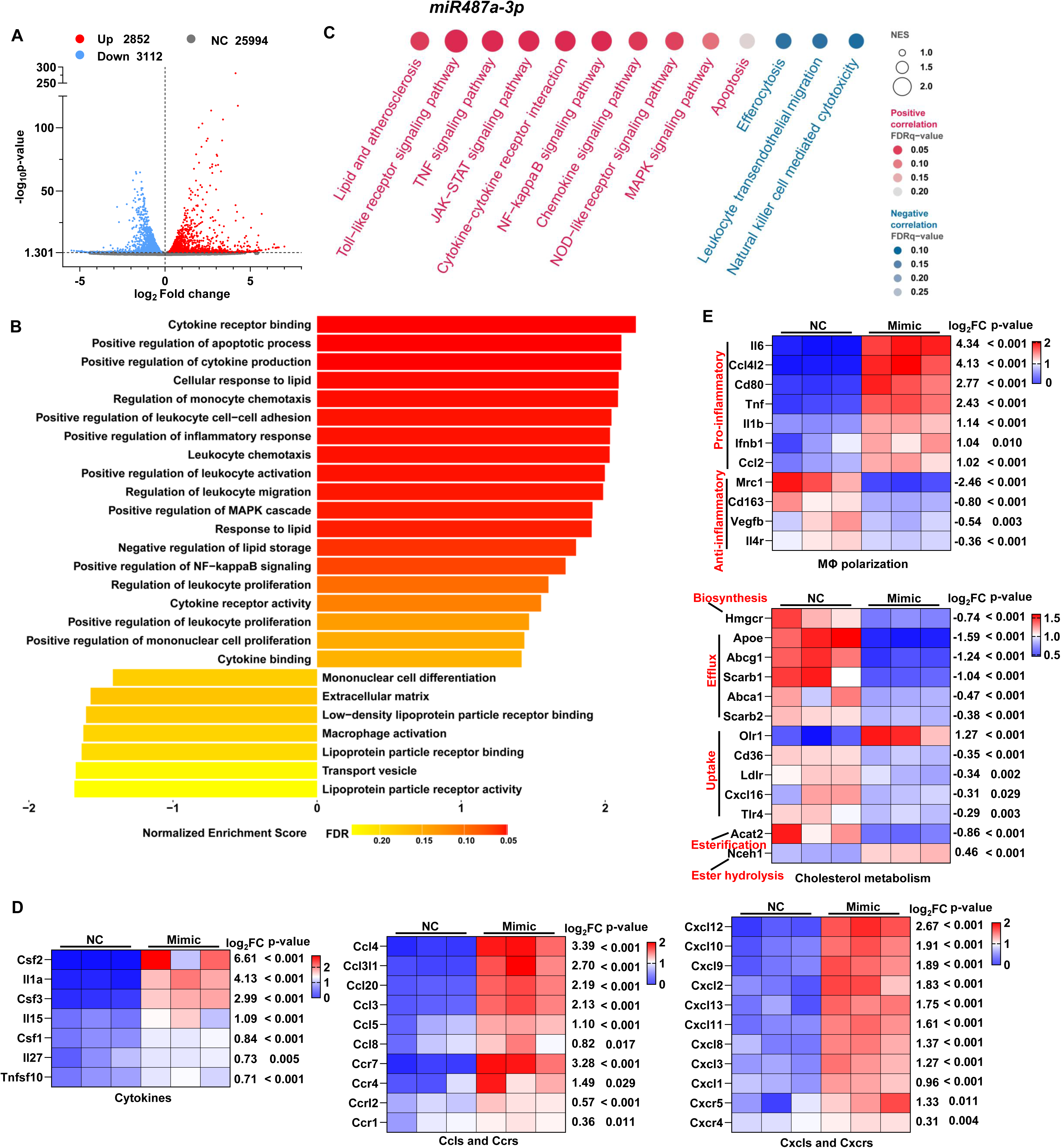
Overexpression of miR487a-3p in THP-1-derived macrophages upregulates signaling related to inflammation and dysregulation of lipid metabolism. **A**, Volcano plot showing differentially expressed genes (DEGs) of THP-1-derived macrophages transfected with miR487a-3p mimic or negative control (NC) for 24 h. **B** and **C**, Gene Set Enrichment Analysis (GSEA) of Gene Ontology (GO) terms (*B*) and Kyoto Encyclopedia of Genes and Genomes (KEGG) pathways (*C*) between NC and miR487a-3p mimic groups. The top enriched GO terms in the molecular function (MF), biological process (BP), and cellular component (CC) categories are shown in (*B*), and the 13 significantly enriched KEGG pathways are shown in (*C*). **D** and **E**, Heatmaps displaying the expression of DEGs related to cytokines, chemokines, and their receptors (*D*), and macrophage polarization and lipid metabolism (*E*). n=3 per group. Abbreviations: CCL, C-C motif chemokine ligand; CCR, C-C chemokine receptor; CXCL, C-X-C motif chemokine ligand; CXCR, C-X-C motif chemokine receptor.

Similarly, miR-6855-3p overexpression induced 1,533 gene upregulation and 1,453 gene downregulation events versus NC (Figure 4A). GSEA-based GO and KEGG analyses revealed enrichment of genes associated with enhanced inflammation, immune dysregulation, and abnormal lipid metabolism (Figure 4B and 4C). miR-6855-3p overexpression upregulated cytokines (Il15, Il27, Csf1, Tnfsf10, Tgfb1) and chemokines (Ccl3, Ccl3l1, Ccl4, Ccl7, Ccl8 and Cxcl8, Cxcl10, Cxcl11, Cxcl12) and their receptors (Ccr1, Ccr7) (Figure 4D; S6D). Pro-inflammatory polarization markers (Il12a, Ifnb1, Cd80, Ccl2, Ccl4l2, Tnf, Il23a) were increased, while the anti-inflammatory marker Il1r2 was downregulated (Figure 4E; S6E). Cholesterol efflux genes (Apoe, Abcg1, Abca5) were suppressed and cholesterol uptake genes (Msr1, Tlr4) were upregulated (Figure 4E; S6E). Extracellular matrix remodeling genes were also affected, with downregulation of Timp1, Timp3 and Mmp1, Mmp2, Mmp3, Mmp8, Mmp9, Mmp13, Mmp14, Mmp17, and upregulation of Mmp13 (Figure S6B). Transcription factor expression was also altered in the miR-6855-3p mimic group compared with NC (Figure S6C and S6F).

**Figure 4.**
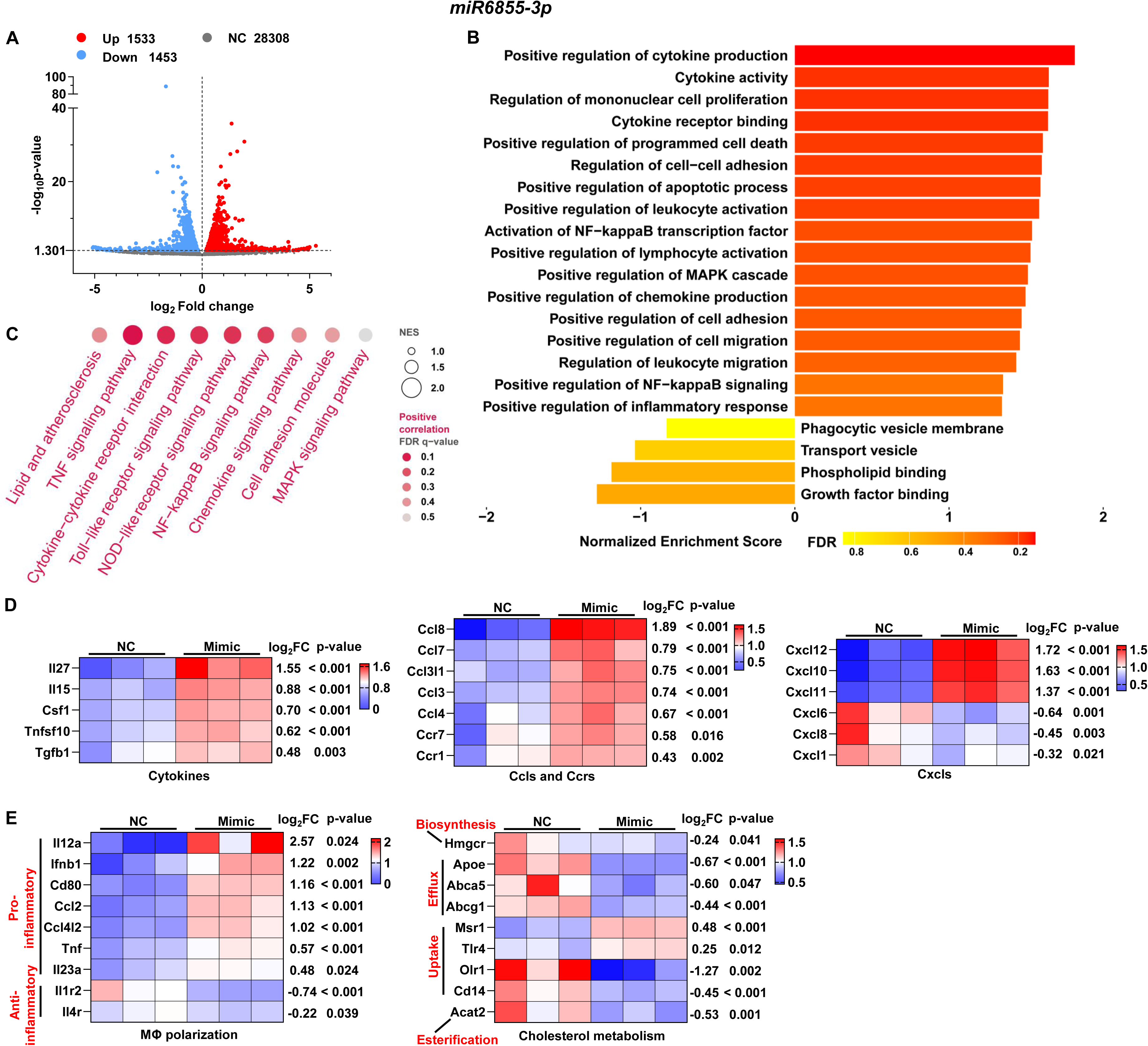
miR6855-3p overexpression drives a shift of gene expressions towards inflammation and lipid accumulation in THP-1-derived macrophages. **A**, Volcano plot showing differentially expressed genes (DEGs) of THP-1-derived macrophages transfected with miR6855-3p mimic or negative control (NC) for 24 h. **B** and **C**, Gene Set Enrichment Analysis (GSEA) of Gene Ontology (GO) terms (*B*) and Kyoto Encyclopedia of Genes and Genomes (KEGG) pathways (*C*) between NC and miR6855-3p mimic groups. The top enriched GO terms in the molecular function (MF), biological process (BP), and cellular component (CC) categories are shown in (*B*), and the 13 significantly enriched KEGG pathways are shown in (*C*). **D** and **E**, Heatmaps displaying the expression of DEGs related to cytokines, chemokines, and their receptors (*D*), and macrophage polarization and lipid metabolism (*E*). n=3 per group. Abbreviations: CCL, C-C motif chemokine ligand; CCR, C-C chemokine receptor; CXCL, C-X-C motif chemokine ligand; CXCR, C-X-C motif chemokine receptor.

Together, these transcriptomic analyses demonstrate that elevated miR487a-3p and miR6855-3p drive broad gene expression programs that converge on pro-inflammatory activation, disrupted lipid metabolism, and extracellular matrix remodeling, constituting hallmarks of pro-atherogenic macrophage polarization.

### miR-487a-3p and miR-6855-3p Promote Macrophage Pro-Inflammatory Polarization, Foam Cell Formation, and Endothelial Injury

We next determined whether the transcriptomic changes induced by miR487a-3p and miR6855-3p translate into functional alterations in macrophage behaviors. miR487a-3p overexpression in THP-1-derived macrophages significantly promoted the secretion of pro-inflammatory cytokines (IL-6, TNF-α, and IL-1β) and chemokines (CCL2, CCL3, CCL4, CXCL8, CXCL9, and CXCL13), while inhibiting TIMP1 and TIMP2 secretion by more than 50%, as measured by antibody array analysis (Figure 5A). Flow cytometry confirmed an increase of the pro-inflammatory marker CD80 and a concomitant decrease of the anti-inflammatory marker CD206 in miR487a-3p-overexpressing macrophages compared with NC (Figure 5B; S7A). To assess the functional consequence of the polarization on the vascular wall, conditioned medium from miR487a-3p-overexpressing macrophages was applied to human aortic endothelial cells (HAECs). It significantly induced HAEC apoptosis compared to treatment with medium from NC macrophages (Figure 5C; S7B), indicating that miR487a-3p-driven macrophage polarization promotes endothelial injury. Furthermore, miR487a-3p overexpression markedly accelerated foam cell formation by enhancing ox-LDL uptake and inhibiting cholesterol efflux (Figure 5D), accompanied by decreased expression of lipid efflux transporters (ABCA1, ABCG1, SCARB1, and APOE) and increased expression of the ox-LDL receptor OLR1 in macrophages (Figure 5E). These key phenotypes were recapitulated in human monocyte-derived macrophages (hMDMs), which similarly exhibited miR-487a-3p overexpression-induced pro-inflammatory polarization, lipid accumulation, and foam cell formation (Figure S7H–K).

**Figure 5.**
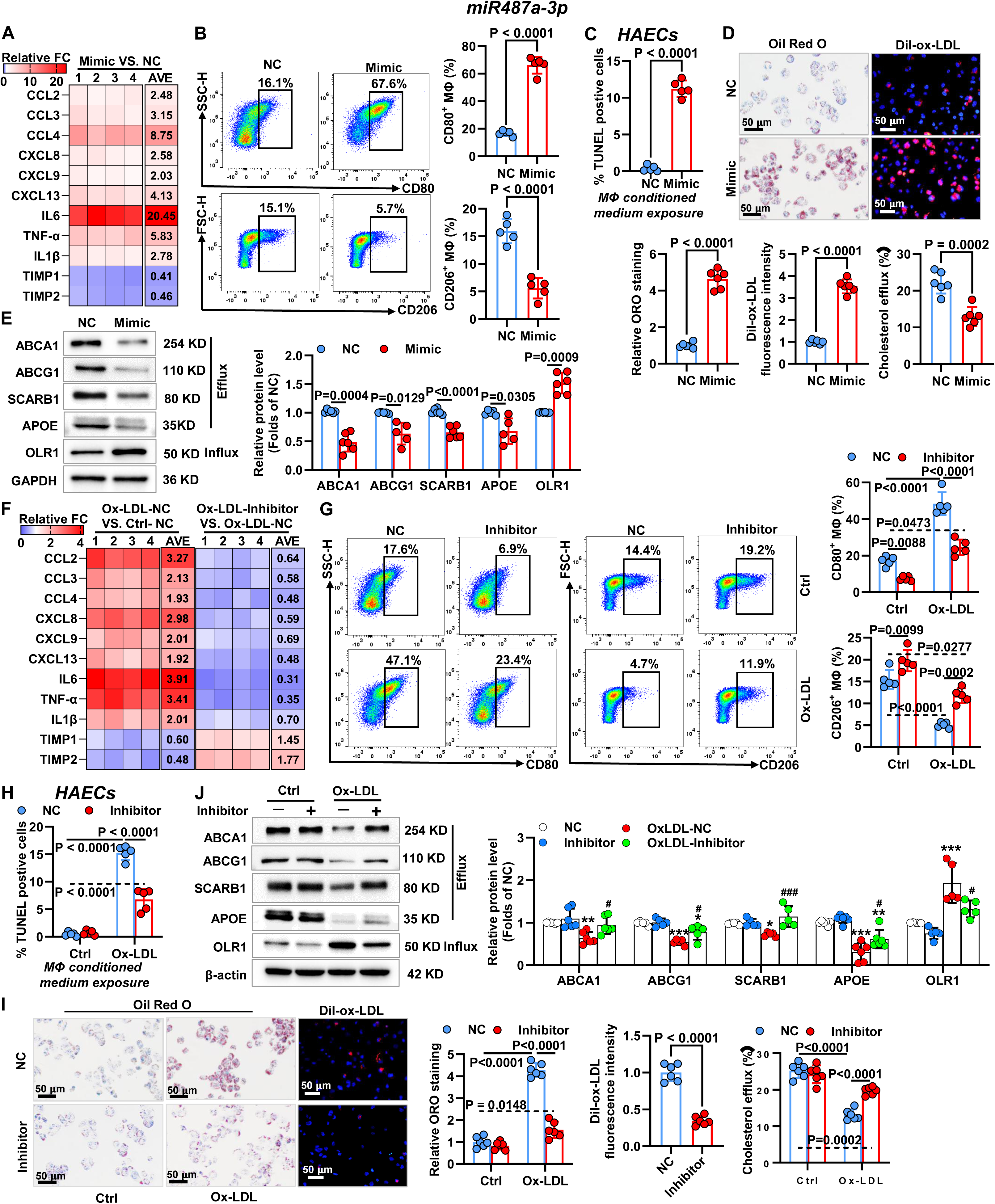
Increases of miR487a-3p mediate macrophages pro-inflammatory polarization and foam cell formation in vitro. **A**, **B**, **D** and **E**, THP-1-derived macrophages were transfected with negative control (NC) or miR487a-3p mimic for 24 h. **F, G, I** and **J**, THP-1-derived macrophages were transfected with NC or miR487a-3p inhibitor for 24 h, followed by incubation with or without 100 μg/mL oxidized low-density lipoprotein (ox-LDL) for an additional 24 h. **C** and **H**, Human aortic endothelial cells (HAECs) were treated with macrophage conditioned medium for 48 h. **A** and **F**, Secretion of soluble factors under the indicated treatment was detected by antibody array analysis. The heatmaps indicating the fold change (FC) compared to NC (***A***, ***F***) or ox-LDL-NC (***F***). The relative expression levels per replicate and the average FC differences were shown (n=4 biologically independent experiments). **B** and **G**, Representative flow cytometry scatter plots and quantification of CD80 and CD206 expression in THP-1-derived macrophages under the indicated conditions (n=5). **C** and **H**, Quantification of TUNEL staining in HAECs (n=5). **D** and **I**, Representative images of Oil Red O staining and Dil-ox-LDL uptake assay with the indicated THP-1-derived macrophages and according group quantification, along with the cholesterol efflux assay (n=6). Scale bar, 50 μm. **E** and **J**, Immunoblotting and quantification of ABCA1, ABCG1, SCARB1, APOE, and OLR1 in THP-1-derived macrophages with the indicated treatment (n=5). GAPDH and β-actin as loading control. Statistics: Unpaired Student’s *t*-test, or Welch’s *t*-test if unequal variance in ***B***-***E*** and ***I***, and 2-way ANOVA with post-hoc Tukey’s multiple comparisons between groups in ***F***-***J***; \**p*<0.05, \*\**p*<0.01, \*\*\**p*<0.001 vs. NC and ^#^*p*<0.05, ^##^*p*<0.01, ^###^*p*<0.001 vs. Ox-LDL-NC in ***J***.

Conversely, inhibition of miR487a-3p in THP-1-derived macrophages and hMDMs significantly attenuated ox-LDL-induced miR487a-3p upregulation (Figure S7C and S7L). miR487a-3p inhibition powerfully suppressed ox-LDL-induced secretion of IL-6, TNF-α, IL-1β, CCL2, CCL3, CCL4, CXCL8, CXCL9, and CXCL13, restored TIMP1 and TIMP2 secretion, and retarded pro-inflammatory polarization in both types of macrophages (Figure 5F and 5G; S7D, S7E, and S7M). Accordingly, endothelial apoptosis induced by conditioned medium from ox-LDL-treated macrophages was significantly blunted by macrophage miR-487a-3p inhibition (Figure 5H; S7F). Ox-LDL-induced foam cell formation was also attenuated by miR-487a-3p inhibition through reduced ox-LDL uptake and restored cholesterol efflux capacity (Figure 5I; S7O), with corresponding recovery of ABCA1, ABCG1, SCARB1, and APOE expression and suppression of OLR1 in two types of macrophages (Figure 5J; S7G and S7N). Together, these data demonstrate that increased miR-487a-3p expression contributes to ox-LDL-induced macrophage inflammation, foam cell formation, and secondary endothelial injury.

miR6855-3p overexpression analogously drove THP-1-derived macrophages to secrete TNF-α and chemokines (CCL2, CCL3, CCL4) while reducing TIMP1 secretion by more than 40% (Figure 6A). Pro-inflammatory polarization was confirmed by upregulation of CD80 and downregulation of the anti-inflammatory marker IL1R2 in both THP-1-derived macrophages and hMDMs (Figure 6B; S8A, S8H, and S8I). Conditioned medium from miR6855-3p-overexpressing macrophages profoundly induced HAEC apoptosis (Figure 6C; S8B). miR6855-3p overexpression also promoted foam cell formation by increasing ox-LDL uptake and impairing cholesterol efflux (Figure 6D; S8K), with corresponding downregulation of ABCG1 and APOE and upregulation of the lipid uptake receptors TLR4 and MSR1 in both types of macrophages (Figure 6E; S8J).

**Figure 6.**
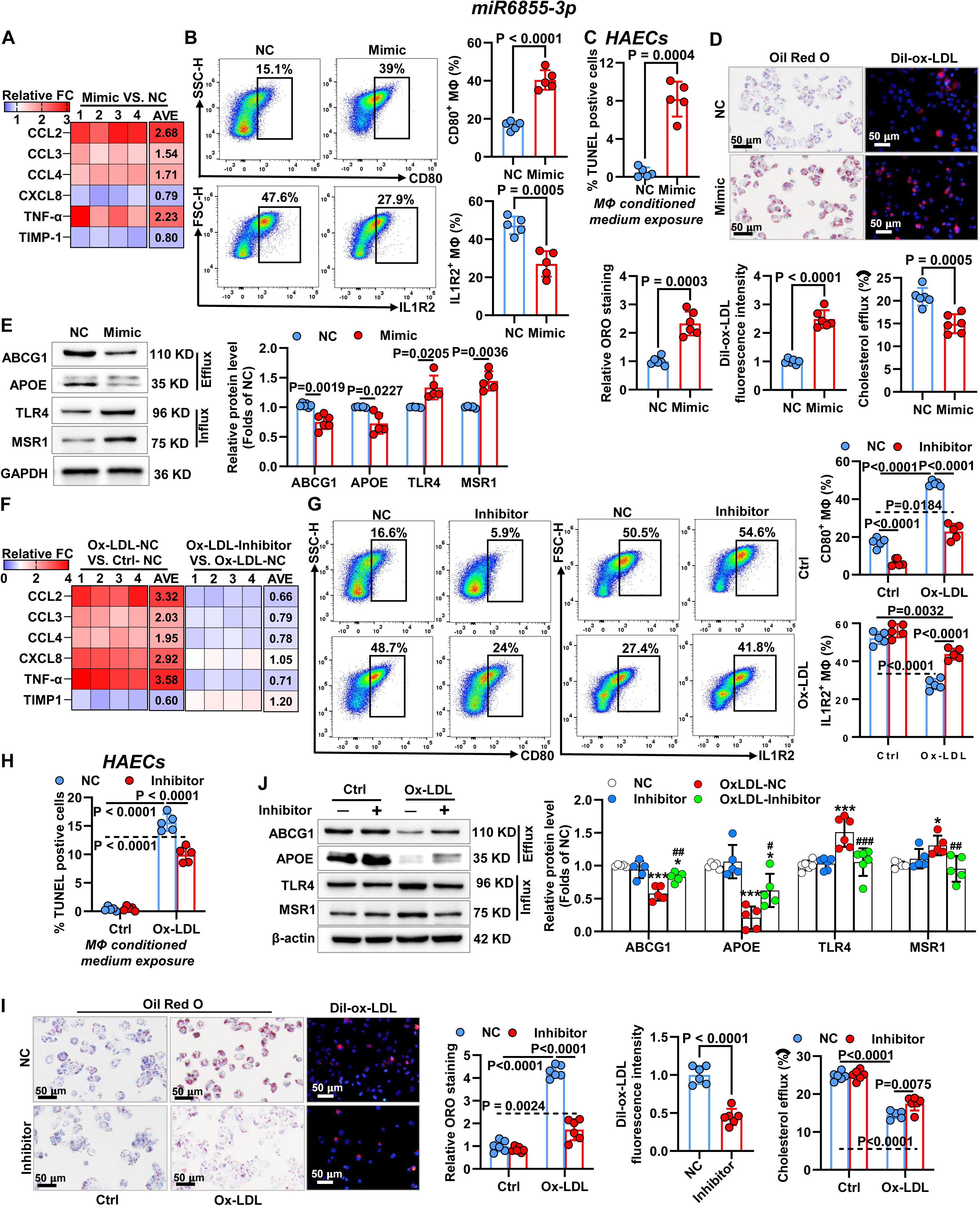
Increases of miR6855-3p facilitate inflammation and lipid accumulation in THP-1-derived macrophages. **A**, **B**, **D** and **E**, THP-1-derived macrophages were transfected with negative control (NC) or miR6855-3p mimic for 24 h. **F, G, I** and **J**, THP-1-derived macrophages were transfected with NC or miR6855-3p inhibitor for 24 h, followed by incubation with or without 100 μg/mL oxidized low-density lipoprotein (ox-LDL) for an additional 24 h. **C** and **H**, Human aortic endothelial cells (HAECs) were treated with macrophage conditioned medium for 48 h. **A** and **F**, Secretion of soluble factors under the indicated treatment was detected by antibody array analysis. The heatmaps indicating the fold change (FC) compared to NC (***A***, ***F***) or ox-LDL-NC (***F***). The relative expression levels per replicate and the average FC differences were shown (n=4 biologically independent experiments). **B** and **G**, Representative flow cytometry scatter plots and quantification of CD80 and IL1R2 expression in THP-1-derived macrophages under the indicated conditions. **C** and **H**, Quantification of TUNEL staining in HAECs. **D** and **I**, Representative images of Oil Red O staining and Dil-ox-LDL uptake assay with the indicated THP-1-derived macrophages and according group quantification, along with the cholesterol efflux assay. Scale bar, 50 μm. **E** and **J**, Immunoblotting and quantification of ABCG1, APOE, TLR4, and MSR1 in THP-1-derived macrophages with the indicated treatment. GAPDH and β-actin as loading control. Data collected from 5-6 independent experiments in ***B***-***E*** and ***G***-***I***. Statistics: Unpaired Student’s *t*-test, or Welch’s *t*-test if unequal variance in ***B***-***E*** and ***I***, and 2-way ANOVA with post-hoc Tukey’s multiple comparisons between groups in ***F***-***J***; \**p*<0.05, \*\*\**p*<0.001 vs. NC and ^#^*p*<0.05, ^##^*p*<0.01, ^###^*p*<0.001 vs. Ox-LDL-NC in ***J***.

Conversely, miR6855-3p inhibition in THP-1-derived macrophages and hMDMs (Figure S8C and S8L) significantly reduced ox-LDL-induced secretion of TNF-α, CCL2, CCL3, and CCL4, restored TIMP1 secretion, and attenuated pro-inflammatory polarization (Figure 6F and 6G; S8D, S8E, and S8M). Endothelial apoptosis induced by conditioned medium from ox-LDL-treated macrophages was correspondingly reduced by miR-6855-3p inhibition (Figure 6H; S8F). Furthermore, miR6855-3p inhibition suppressed ox-LDL-induced foam cell formation by reducing ox-LDL uptake and restoring cholesterol efflux (Figure 6I; S8O), with reversal of ox-LDL-induced ABCG1 and APOE downregulation and TLR4 and MSR1 upregulation in both types of macrophages (Figure 6J; S8G and S8N). Taken together, these results demonstrate that elevated miR-6855-3p expression mediates ox-LDL-induced macrophage pro-inflammatory polarization, lipid accumulation, and consequent endothelial injury.

### miR-487a-3p and miR-6855-3p Directly Target CPE and RRM2, Respectively, to Promote Macrophage Pro-Inflammatory Polarization and Foam Cell Formation

To identify the direct targets mediating the pro-atherogenic effects of these miRNAs, we performed mRNA-seq on peripheral monocytes from CAD patients and HCs (Table 5) and identified 605 DEGs (CAD vs. HC) (Figure 7A). GO and KEGG analyses revealed enrichment of genes associated with inflammation and lipid metabolism (Figure S9A and S9B). For each miRNA, we intersected TargetScan-predicted targets with downregulated DEGs from THP-1-derived macrophages (mimic vs. NC) and from peripheral monocytes (CAD vs. HC). Venn analysis of macrophage DEGs and predicted targets identified 13 candidates for miR487a-3p (Figure S10A) and 142 candidates for miR6855-3p (Figure S11A); the top eight candidates for each were validated by RT-qPCR (Figures S10C and S11C). Intersection of monocyte DEGs with predicted targets yielded three candidates for miR487a-3p (Figure S10B) and 42 for miR6855-3p (Figure S11B). A three-way Venn analysis combining macrophage DEGs, monocyte DEGs, and predicted targets identified CPE (carboxypeptidase E) as the solely convergent candidate for miR487a-3p (Figure 7B), ranking highest in both intersections and showing the most significant expression change in miR487a-3p mimic-transfected versus NC macrophages (Figure S10A-S10C). For miR6855-3p, three candidates were identified in the same way, among which RRM2 (ribonucleotide reductase subunit M2) ranked highest in both intersections and exhibited the most significant expression change (Figure 7B; S11A-S11C).

**Figure 7.**
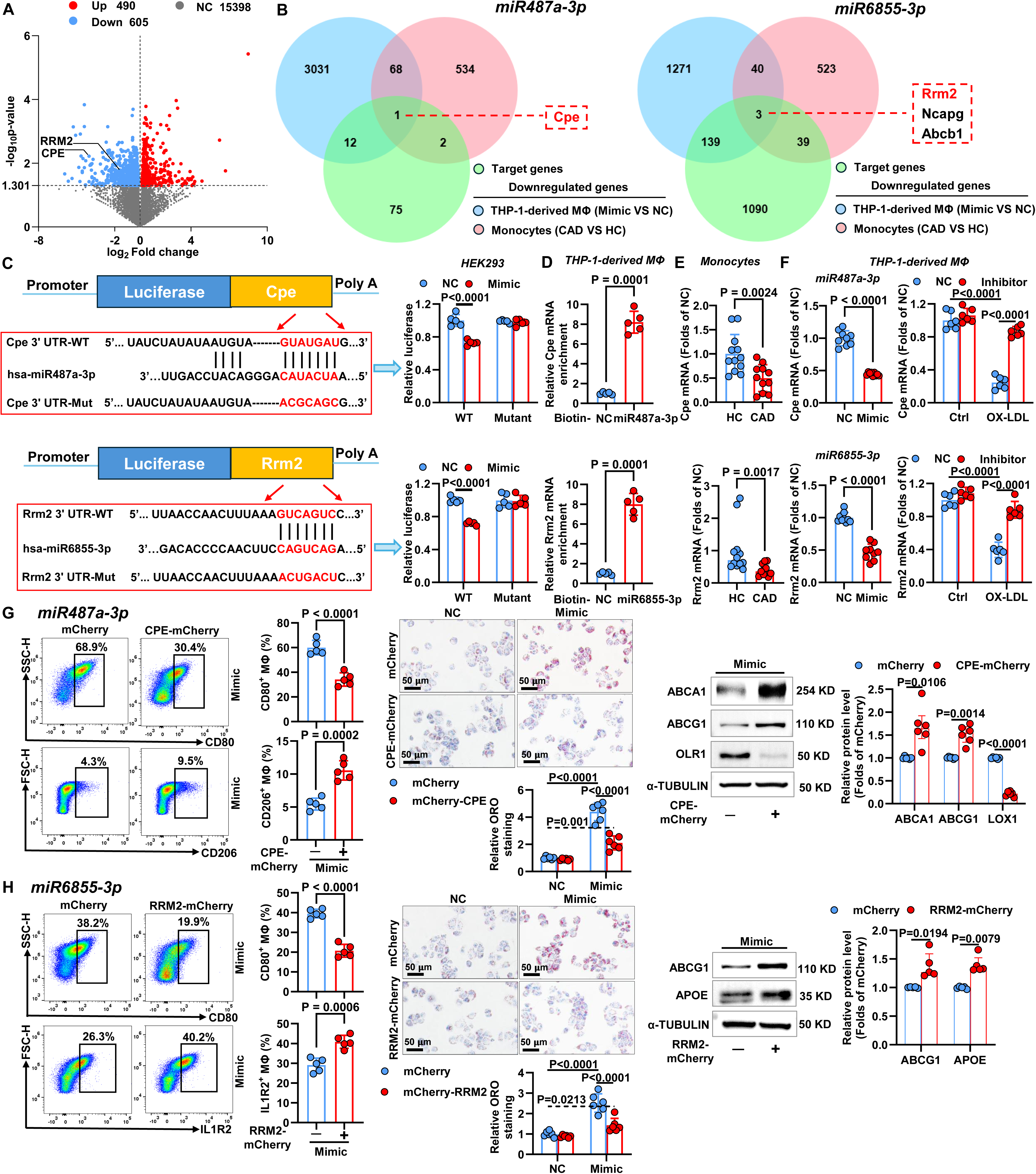
miR487a-3p and miR6855-3p target CPE and RRM2 respectively, promoting macrophages pro-inflammatory polarization and lipid accumulation. **A**, Volcano plot showing differentially expressed genes (DEGs) in peripheral monocytes from healthy controls (HCs, n=10) and coronary artery disease (CAD) patients (n=9). **B**, Venn diagrams depicting the overlap between TargetScan-predicted targets of miR487a-3p or miR6855-3p and downregulated genes in THP-1-derived macrophages (miR487a-3p or miR6855-3p mimic vs. negative control, NC) and in peripheral monocytes (CAD vs. HC). **C**, Schematic representation of the predicted binding sites for miR487a-3p and miR6855-3p, and the mutated binding sites in the 3′ UTR of Cpe and Rrm2. HEK-293T cells were co-transfected with dual-luciferase reporter vectors containing wild-type (WT) or mutant (MT) Cpe or Rrm2 3′ UTR, and with miR487a-3p mimic or 6855-3p mimic or NC. Relative luciferase activity was quantified. **D**, Relative levels of Cpe or Rrm2 recruited by Biotin-miR-487a-3p, or Biotin-miR-6855-3p or Biotin-miR-NC probes were measured by miRNA pull-down assay in THP-1-derived macrophages. **E**, The mRNA expressions of Cpe and Rrm2 in peripheral monocytes from HCs (n=12) and CAD patients (n=11). **F**, The mRNA expressions of CPE or RRM2 in THP-1-derived macrophages transfected with NC or miR487a-3p mimic or 6855-3p mimic for 24 h, or transfected with NC or miR487a-3p inhibitor or miR6855-3p inhibitor for 24 h followed by treatment with control (Ctrl) or 100 μg/mL oxidized low-density lipoprotein (ox-LDL) for 24 h. **G** and **H**, THP-1-derived macrophages were infected with lentiviral vectors containing mCherry-CPE (*G*) or mCherry-RRM2 (*H*) or mCherry, and further transfected with NC or miR487a-3p mimic (*G*) or 6855-3p mimic (*H*) for 24 h. Left: Representative flow cytometry scatter plots and quantification of CD80, CD206 (*G*) and IL1R2 (*H*) expression frequency. Middle: Representative images of Oil Red O staining and quantification. Right: Immunoblotting and quantitation of ABCG1, ABCA1 (*G*), OLR1 (*G*) and APOE (*H*), and GAPDH as loading control. Data collected from 5-6 independent experiments in *C*, *D* and *F*-*H*. Statistics: 2-way ANOVA with post-hoc Tukey’s multiple comparisons between groups in *C* and *F*. Unpaired Student’s *t*-test in *E*-*H*, Welch’s *t*-test in *D* and *F*-*H*, and Mann-Whitney *U* test in *E* and *H*.

**Table 5.** Demographic data of the health controls (HCs) and coronary artery disease (CAD) patients for monocyte mRNA sequencing.

|  | **HC (n=10)** | **CAD (n=9)** | **P- value** |
| --- | --- | --- | --- |
| **Age (years)** | 60.70 ± 5.98 | 64.00 ± 5.79 | 0.2395 |
| **Male n. (%)** | 5 (50.00%) | 5 (55.56%) | >0.9999 |
| **Body mass index (kg/m2)** | 22.35 ± 2.13 | 24.04 ± 2.73 | 0.1494 |
| **Smoking n. (%)** | 1 (10.00%) | 5 (55.56%) | 0.0573 |
| **Drinking n. (%)** | 1 (10.00%) | 4 (44.44%) | 0.1409 |
| **Hypertension n. (%)** | 0 (0%) | 0 (0%) | >0.9999 |
| **Diabetes mellitus n. (%)** | 0 (0%) | 0 (0%) | >0.9999 |
| **Statin use n. (%)** | 7 (70.00%) | 9 (100.00%) | 0.2105 |
| **ACEI/ARB use n. (%)** | 0 (0%) | 3 (33.33%) | 0.0867 |
| **β-blocker use n. (%)** | 2 (20.00%) | 3 (33.33%) | 0.6285 |
| **White blood cells (x10<sup>9</sup>/L)** | 5.46 (5.30 - 7.25) | 6.56 (5.90 - 7.81) | 0.2342 |
| **Neutrophils (x10<sup>9</sup>/L)** | 3.86 ± 1.15 | 4.37 ± 1.17 | 0.3534 |
| **Lymphocytes (x10<sup>9</sup>/L)** | 1.84 ± 0.55 | 1.68 ± 0.49 | 0.539 |
| **Monocytes (x10<sup>9</sup>/L)** | 0.46 ± 0.09 | 0.49 ± 0.08 | 0.4693 |
| **Alanine aminotransferase (U/L)** | 11.00 (10.00 - 25.00) | 17.00 (10.00 - 22.00) | 0.9222 |
| **Aspartate aminotransferase (U/L)** | 17.60 ± 3.31 | 20.11 ± 5.44 | 0.2352 |
| **Total bilirubin (μmol/L)** | 12.03 ± 4.19 | 11.88 ± 3.37 | 0.932 |
| **Blood urea nitrogen (mmol/L)** | 6.23 ± 0.81 | 6.02 ± 0.63 | 0.5305 |
| **Creatinine (μmol/L)** | 68.41 ± 13.77 | 73.02 ± 11.60 | 0.4436 |
| **Total cholesterol (mmol/L)** | 4.28 ± 0.55 | 5.26 ± 0.60 | **0.0017** |
| **Triglycerides (mmol/L)** | 0.81 (0.61 - 1.26) | 1.40 (0.89 - 1.60) | 0.0751 |
| **HDL-C (mmol/L)** | 1.45 ± 0.33 | 1.27 ± 0.31 | 0.2542 |
| **LDL-C (mmol/L)** | 2.18 ± 0.60 | 2.93 ± 0.55 | **0.012** |
| **SdLDL-C (mmol/L)** | 0.66 ± 0.22 | 1.14 ± 0.32 | **0.0014** |
| **Fasting blood glucose (mmol/L)** | 5.20 ± 0.37 | 5.50 ± 0.78 | 0.3065 |
| **Left ventricular ejection fraction (%)** | 65.70 ± 2.79 | 63.56 ± 4.95 | 0.2546 |

Data are expressed as mean ± SD or median with interquartile if non-normal distribution, and n (%). ACEI, angiotension converting enzyme inhibitor; ARB, angiotensin receptor blocker; HDL-C, high-density lipoprotein cholesterol; LDL-C, low-density lipoprotein cholesterol; SdLDL-C, small dense low-density lipoprotein cholesterol.

Dual-luciferase assays in human embryonic kidney (HEK)-293 cells and THP-1-derived macrophages verified the predicted binding of miR487a-3p to the CPE 3′ UTR and miR6855-3p to the RRM2 3′ UTR (Figure 7C). miR487a-3p mimic significantly reduced the luciferase activity of the wild-type (WT) CPE 3′ UTR without affecting the mutant in both cell types, while miR-487a-3p inhibitor restored the luciferase activity of the WT CPE 3′ UTR that was suppressed by ox-LDL-induced miR487a-3p upregulation in macrophages; neither ox-LDL nor the inhibitor affected the mutant (Figure 7C; S12A and S13A). Similar results were obtained for the binding of miR-6855-3p and the RRM2 3′ UTR (Figure 7C; S12A and S13E). miRNA pulldown assays further confirmed the direct physical interaction: biotin-labeled miR487a-3p markedly enriched CPE mRNA and biotin-labeled miR6855-3p robustly enriched RRM2 mRNA compared with biotin-miR-NC (Figure 7D). Consistently, Cpe and Rrm2 mRNA levels were decreased in circulating monocytes of CAD patients compared with HCs (Figure 7E; Table S4). In THP-1-derived macrophages, miR487a-3p and miR6855-3p mimics significantly reduced CPE and RRM2 mRNA and protein expression, respectively, while ox-LDL treatment alone suppressed both CPE and RRM2, an effect reversed by the corresponding miRNA inhibitors (Figure 7F; S13B and S13F).

CPE is a processing enzyme with established links to metabolic and cardiovascular diseases,^27–29^ and its deficiency has been implicated in inflammatory pathologies. To determine whether CPE mediates the functional effects of miR487a-3p, we performed rescue experiments in THP-1-derived macrophages. CPE overexpression via lentiviral infection (Figure S12B and S13C) partially reversed miR487a-3p-induced macrophage pro-inflammatory polarization and foam cell formation (Figure 7G), with decreased expression of inflammatory cytokines and chemokines (Ccl3, Ccl4, Cxcl8, Cxcl13), pro-inflammatory markers (CD80, Ccl2, Il6, Tnf), and the lipid uptake receptor OLR1, and increased expression of anti-inflammatory markers (CD206, Cd163) and cholesterol efflux transporters (ABCA1, ABCG1) (Figure 7G; S13D). RRM2 is essential for nucleotide metabolism and maintaining dNTP pools,^30^ and its inhibition has been associated with promoting macrophage pro-inflammatory polarization.^30–32^ Analogously, RRM2 overexpression (Figure S12B and S13G) substantially abrogated miR6855-3p-induced pro-inflammatory polarization and foam cell formation (Figure 7H), suppressing inflammatory chemokines (Ccl3, Ccl4, Cxcl8) and pro-inflammatory markers (CD80, Ccl2, Tnf) while enhancing the anti-inflammatory marker IL1R2 and cholesterol efflux transporters (Abcg1, Apoe) (Figure 7H; S13G). Together, these results demonstrate that miR487a-3p and miR6855-3p promote macrophage pro-atherogenic transformation by directly targeting CPE and RRM2, respectively.

### Myeloid-Specific Deletion of CPE or RRM2 Accelerates Atherosclerosis in Hypercholesterolemic Mice

To validate the pathogenic role of the miR487a-3p-CPE and miR6855-3p-RRM2 axes in vivo, we generated mice with monocyte/macrophage-specific deletion of Cpe or Rrm2 using the Lyz2^CreERT^^2^ system, thereby mimicking the functional consequence of elevated miRNA levels observed in CAD patients. Direct miRNA inhibition in mice is precluded by the absence of miR-487a-3p and miR-6855-3p in the murine genome. This approach is feasible because, among myeloid cells, CPE and RRM2 are expressed primarily in monocytes and macrophages, with negligible expression in neutrophils in low-density lipoprotein receptor (Ldlr)^-/-^ mice on either chow diet (CD) or high-fat diet (HFD) (data not shown). Consistently, CPE^+^ or RRM2^+^ neutrophils constituted <1% of total neutrophils in Cpe^f/f^ or Rrm2^f/f^ mice (Figure S14A). All the floxed and Lyz2^CreERT^^2^ mice received AAV-mediated proprotein convertase subtilisin/ kexin type (PCSK) 9 overexpression to induce a Ldlr knockout phenotype and hypercholesterolemia^33^ and were fed CD or HFD containing tamoxifen for 14 weeks (Figure 8A). Tamoxifen treatment extensively depleted CPE^+^ or RRM2^+^ monocytes and macrophages without affecting CPE or RRM2 expression in Cpe^f/f^ or Rrm2^f/f^ controls (Figure S14A and S14B).

**Figure 8.**
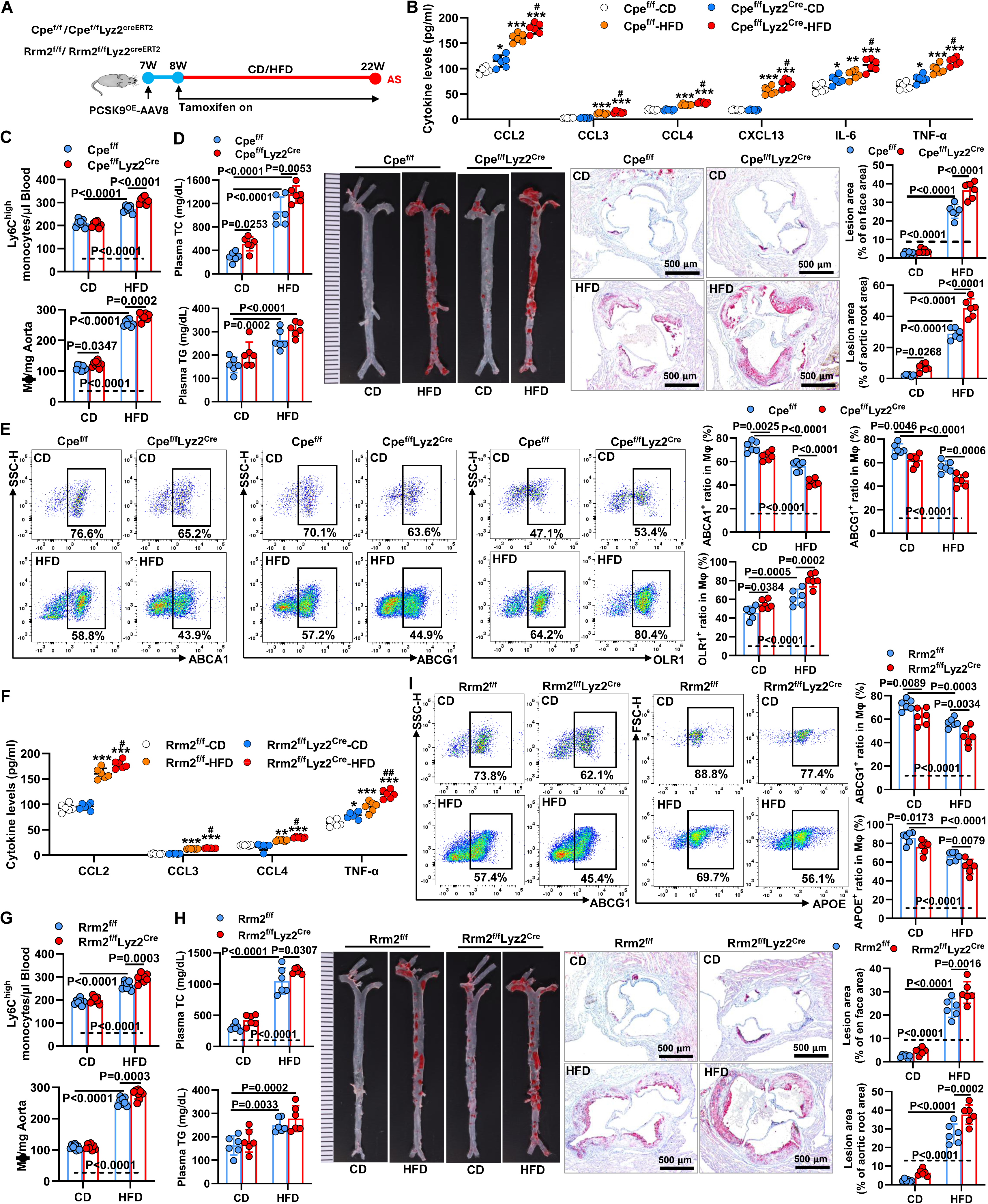
Myeloid-specific CPE or RRM2 depletion aggravates inflammation and promotes atherosclerosis development in PCSK9 overexpression mice on high-fat diet (HFD). **A**, Protocol for myeloid-specific CPE or RRM2 ablation in Cpe^f/f^- or Rrm2^f/f^-Lyz2^CreERT2^ mice (Cpe^f/f^ or Rrm2^f/f^ littermates as controls) with tamoxifen administration in chow diet (CD) or HFD from 8 to 22 weeks after 1 week of PCSK9^OE^-AAV8 infection. **B** and **F**, Plasma levels of CC chemokine ligand (CCL) 2, CCL3, CCL4, C-X-C Motif Chemokine Ligand (CXCL) 8, CXCL13 (*B*), interleukin (IL) 6 (*B*) and tumor necrosis factor (TNF)-α in myeloid-specific CPE (*B*) or RRM2 (*F*) deletion mice (n=6 per group). **C** and **G**, Quantitation of blood Ly6C^high^ monocytes and aortic macrophages in myeloid-specific CPE (*C*) or RRM2 (*G*) knockout mice (n=8 per group). **D** and **H**, Plasma levels of total cholesterol (TC) and triglycerides (TG) (Left), and representative Oil Red O staining images of whole aortas and aortic roots (Middle) and corresponding quantification (Right) in mice harboring specific myeloid cell deficiency of CPE (*D*) or RRM2 (*H*) (n=6 per group). **E** and **I**, Flow cytometry plots and quantitation of ABCG1, ABCA1 (*E*), OLR1 (*E*) and APOE (*I*) expressions in aortic macrophages from mice harboring myeloid-specific deletion of CPE (*E*) or RRM2 (*I*) (n=6 per group). Statistics: 2-way ANOVA with post-hoc Tukey’s multiple comparisons between groups in *B*-*I*; \**p*<0.05, \*\**p*<0.01, \*\*\**p*<0.001 vs. Cpe^f/f^-CD (*B*) or Rrm2^f/f^-CD (*F*), and ^#^*p*<0.05, ^##^*p*<0.01, ^###^*p*<0.001 vs. Cpe^f/f^Lyz2^Cre^-CD (*B*) or Rrm2^f/f^Lyz2^Cre^-CD (*F*) in *B* and *F*.

In Cpe^f/f^ mice, HFD increased plasma levels of inflammatory cytokines and chemokines (CCL2, CCL3, CCL4, CXCL13, IL-6, and TNF-α), circulating Ly6C^high^ monocytes, and aortic macrophages, all of which were further elevated in Cpe^f/f^Lyz2^Cre^-HFD mice (Figure 8B and 8C). Cpe^f/f^Lyz2^Cre^-CD mice also exhibited increased plasma CCL2, IL-6, and TNF-α levels and aortic macrophages compared with Cpe^f/f^-CD mice, whereas peripheral Ly6C^low^ monocyte numbers were unchanged across all groups (Figure S14A). In Cpe^f/f^ mice, HFD induced atherosclerotic plaque formation in the whole aorta and aortic root, as well as elevated plasma TC and TG levels, which were further aggravated in Cpe^f/f^Lyz2^Cre^-HFD mice, except for plasma TG (Figure 8D). Notably, plasma TC levels and aortic root lesion area were also increased in Cpe^f/f^Lyz2^Cre^ mice on CD. As opposed to Cpe^f/f^ mice, the HFD-induced suppression of ABCA1 and ABCG1 and upregulation of OLR1 in aortic macrophages were exacerbated by myeloid-specific CPE ablation, which also impaired ABCA1 and ABCG1 expression and increased OLR1 in CD-fed mice (Figure 8E).

Similarly, in Rrm2^f/f^ mice, HFD elevated plasma inflammatory cytokines and chemokines (CCL2, CCL3, CCL4, and TNF-α), TC and TG, peripheral Ly6C^high^ monocytes, and aortic macrophages, and promoted atherosclerotic lesion formation, all of which were further intensified in Rrm2^f/f^Lyz2^Cre^-HFD mice except plasma TG (Figure 8F–H). Neither HFD nor myeloid-specific RRM2 depletion affected circulating Ly6C^low^ monocyte numbers (Figure S14A). HFD-induced downregulation of ABCG1 and APOE in aortic macrophages was further exacerbated by RRM2 deficiency, which also reduced ABCG1 and APOE expression and increased plasma TNF-α in CD-fed mice (Figure 8F and I).

Together, these in vivo data demonstrate that monocyte/macrophage-specific deletion of Cpe or Rrm2 accelerates atherosclerosis in hypercholesterolemic mice by promoting macrophage inflammatory responses and lipid accumulation, directly recapitulating the in vitro phenotypes induced by miR487a-3p and miR6855-3p overexpression. These findings establish CPE and RRM2 as the key downstream targets of these miRNAs in atherogenesis.

## DISCUSSION

We have identified miR487a-3p and miR6855-3p as novel biomarkers and pathogenic mediators in CAD. Both miRNAs were upregulated in peripheral monocytes, PBMCs, and coronary plaque macrophages of CAD patients, with their plasma levels correlating with blood lipids and Gensini score (AUC≈0.83 for each), supporting their diagnostic and prognostic value. Their transcription was driven by ox-LDL via KLF5 and IRF1, respectively. Notably, both miRNAs were predominantly expressed in coronary arterial macrophages, and the abundance of miR487a-3p⁺ and miR6855-3p⁺ macrophages was positively associated with plaque area, identifying these macrophage-derived miRNAs as potential therapeutic targets for CAD. In vitro, both miRNAs promoted macrophage pro-inflammatory polarization, foam cell formation, and endothelial injury in vitro. These effects were mediated through direct targeting of CPE and RRM2, as confirmed by dual-luciferase assays, miRNA pulldown, and functional rescue experiments. Importantly, myeloid-specific deletion of Cpe or Rrm2 in hypercholesterolemic mice recapitulated the pro-atherogenic phenotypes, establishing the miRNA-CPE/RRM2 axes as drivers of AS progression in vivo.

miR487a-3p has been previously implicated in tumor suppression, autoimmune pathology, and other diseases through multiple reported targets (Table S5). Notably, none of the binding between miR487a-3p and these genes was reported in macrophages (Table S5). Among its 12 known targets, only four (Akip1, Wnt5a, Ccnd1, and Sox9) were downregulated in miR487a-3p mimic-transfected versus NC macrophages according to our mRNA-seq data, and none of these four genes showed differential expression in peripheral monocytes between CAD patients and controls (Figure S10D), indicating that they are unlikely to mediate miR487a-3p function in the context of CAD. In addition to these known functions, our study reveals a pro-inflammatory and pro-atherogenic role for miR487a-3p in macrophages, where it targets Cpe to promote inflammation and lipid dysregulation, thereby accelerating AS. Similarly, miR-6855-3p was previously reported to promote antioxidant protection in breast cancer cells via non-canonical translational activation of PRDX5A.^34^ Although PRDX5A is expressed in macrophages and implicated in redox-sensitive inflammatory signaling^35^, PRDX5A expression did not differ between miR-6855-3p mimic and NC groups in macrophages, nor between CAD and HC groups in peripheral monocytes (Figure S11D). Instead, elevated miR6855-3p promoted macrophage pro-inflammatory polarization and foam cell formation through canonical 3′UTR targeting of RRM2. These findings underscore the context-dependent pleiotropy of miRNAs, wherein a single miRNA can drive divergent, even opposing, outcomes depending on cell type, target gene repertoire, and disease milieu.

The elevated plasma levels of miR487a-3p and miR6855-3p likely reflect their release from activated monocytes and macrophages, as demonstrated by our in vitro secretion assays. Circulating miRNAs are remarkably stable due to their association with extracellular vesicles, Argonaute proteins, and lipoproteins,^1,16,36^ and can mediate intercellular communication.^37,38^ The positive correlation of both miRNAs with the Gensini score and their robust ROC performance support their utility as minimally invasive biomarkers for CAD diagnosis and prognosis. Importantly, multivariate logistic regression analysis demonstrated that both miRNAs remained significant independent predictors of CAD after adjustment for TC, LDL-C, and sdLDL-C, whereas the lipid parameters themselves lost significance. This may reflect the higher prevalence of cholesterol-lowering medication use in the CAD group (Table 2), which reduces circulating lipid levels without necessarily reversing the miRNA upregulation initially driven by hypercholesterolemia. These findings support the potential utility of these miRNAs as stable biomarkers of atherogenic burden that complement traditional lipid profiling.

Monocytes infiltrate the arterial intima in response to endothelial injury, differentiate into macrophages, and drive plaque progression through local proliferation.^39,40^ We found that miR-487a-3p⁺ and miR-6855-3p⁺ macrophages constituted ∼60% of total plaque macrophages in CAD patients, and their abundance positively correlated with lesion area. Based on our functional data, we postulate that aberrant upregulation of these miRNAs in circulating monocytes promotes their pro-inflammatory activation and recruitment into plaques, while sustained miRNA expression in lesional macrophages drives foam cell formation and inflammatory cytokine secretion, amplifying atherogenesis. Inhibition of either miRNA substantially attenuated ox-LDL-induced inflammatory responses and lipid accumulation, supporting their potential as therapeutic targets. Additionally, both miRNAs are lipid-inducible, with their transcription mediated by KLF5 and IRF1 respectively, providing a mechanistic link between hyperlipidemia and miRNA-driven macrophage activation in CAD.

CPE, a prohormone processing enzyme linked to metabolic and cardiovascular diseases,^27–29^ was suppressed by miR487a-3p in monocytes and macrophages. CPE has been shown to exert anti-inflammatory effects in the intestine by modulating cytokine levels and myeloid cell migration,^41^ and its overexpression protects cardiomyocytes from ischemia/hypoxia injury.^27^ Consistent with these observations, CPE overexpression mitigated miR487a-3p-induced inflammation and restored lipid metabolic homeostasis in our study. Importantly, myeloid-specific CPE deficiency exacerbated inflammation, impaired cholesterol efflux transporter expression, and accelerated AS in hypercholesterolemic mice, indicating a protective role for CPE in atherogenesis. RRM2, essential for dNTP synthesis and genome maintenance,^30^ has been reported to regulate macrophage polarization, with its inhibition promoting pro-inflammatory phenotypes.^31,32^ We found that miR6855-3p suppressed RRM2 expression, and RRM2 overexpression reversed miR6855-3p-induced pro-inflammatory polarization and foam cell formation. Conversely, myeloid-specific RRM2 deficiency exacerbated inflammation and AS in vivo. These results establish RRM2 as a critical regulator of macrophage lipid metabolism and inflammatory homeostasis. Together, these findings identify CPE and RRM2 as functionally convergent downstream effectors of miR-487a3p and miR6855-3p, respectively, whose suppression coordinates macrophage inflammatory activation and lipid metabolic dysfunction in AS.

Our results must be considered in the context of potential study limitations. First, validation in larger, independent cohorts is needed to confirm the clinical utility of these miRNAs as biomarkers and to develop robust multi-variable prediction models. Second, MiRNA mimic transfection achieved supraphysiological levels, although key findings were corroborated by inhibitor rescue and in vivo target gene deletion. Last, direct in vivo miRNA inhibition would further strengthen the translational relevance; this represents an important future direction that would require humanized miRNA mouse models.

In summary, miR-487a-3p and miR-6855-3p, through targeting CPE and RRM2 respectively, drive macrophage pro-inflammatory responses and lipid metabolic dysregulation in AS. Their release into the circulation, combined with their correlation with CAD severity, establishes these miRNAs as novel biomarkers for CAD diagnosis and prognosis, while their pathogenic mechanisms identify them as potential therapeutic targets.

## Supplemental Material

Detailed Methods

Supplemental Figures 1–14

Supplemental Tables 1–7

Major Resources Table

References 42–49

## NONSTANDARD ABBREVIATIONS AND ACRONYMS

CAD: coronary artery disease
AS: atherosclerosis
PBMC: peripheral blood mononuclear cell
hMDM: human monocyte-derived macrophage
miRNA: microRNA
ox-LDL: oxidized low-density lipoprotein
HDL-C: high-density lipoprotein cholesterol
LDL-C: low-density lipoprotein cholesterol
sdLDL-C: small dense low-density lipoprotein cholesterol
ABCA1: ATP binding cassette subfamily A member 1
ABCG1: ATP binding cassette subfamily G member 1
SCARB1: scavenger receptor class B member 1
APOE: apolipoprotein E
OLR1: oxidized low density lipoprotein receptor 1
TLR4: toll like receptor 4
MSR1: macrophage scavenger receptor 1
CPE: carboxypeptidase E
RRM2: ribonucleotide reductase regulatory subunit M2
KLF5: Krüppel-like factor 5
IRF1: interferon regulatory factor 1

## REFERENCES

1. Makarova J, Turchinovich A, Shkurnikov M, Tonevitsky A. Extracellular miRNAs and Cell-Cell Communication: Problems and Prospects. Trends Biochem Sci. 2021;46:640–651. doi: 10.1016/j.tibs.2021.01.007

2. Choudhury RR, Gupta H, Bhushan S, Singh A, Roy A, Saini N. Role of miR-128-3p and miR-195-5p as biomarkers of coronary artery disease in Indians: a pilot study. Scientific Reports. 2024;14. doi: 10.1038/s41598-024-61077-4

3. Ali Sheikh MS, Alduraywish A, Almaeen A, Alruwali M, Alruwaili R, Alomair BM, Salma U, Hedeab GM, Bugti N, A.M.Abdulhabeeb I, et al. Therapeutic Value of miRNAs in Coronary Artery Disease. Oxidative Medicine and Cellular Longevity. 2021;2021. doi: 10.1155/2021/8853748

4. Kondracki B, Kłoda M, Jusiak-Kłoda A, Kondracka A, Waciński J, Waciński P. MicroRNA Expression in Patients with Coronary Artery Disease and Hypertension—A Systematic Review. International Journal of Molecular Sciences. 2024;25. doi: 10.3390/ijms25126430

5. Ghafouri-Fard S, Gholipour M, Taheri M. Role of MicroRNAs in the Pathogenesis of Coronary Artery Disease. Frontiers in Cardiovascular Medicine. 2021;8. doi: 10.3389/fcvm.2021.632392

6. Zaman S, Wasfy JH, Kapil V, Ziaeian B, Parsonage WA, Sriswasdi S, Chico TJA, Capodanno D, Colleran R, Sutton NR, et al. The Lancet Commission on rethinking coronary artery disease: moving from ischaemia to atheroma. Lancet. 2025;405:1264–1312. doi: 10.1016/s0140-6736(25)00055-8

7. Jiang Y, Zhao Y, Li Z-y, Chen S, Fang F, Cai J-h. Potential roles of microRNAs and long noncoding RNAs as diagnostic, prognostic and therapeutic biomarkers in coronary artery disease. International Journal of Cardiology. 2023;384:90–99. doi: 10.1016/j.ijcard.2023.03.067

8. Moore KJ, Sheedy FJ, Fisher EA. Macrophages in atherosclerosis: a dynamic balance. Nature Reviews Immunology. 2013;13:709–721. doi: 10.1038/nri3520

9. Theofilis P, Oikonomou E, Tsioufis K, Tousoulis D. The Role of Macrophages in Atherosclerosis: Pathophysiologic Mechanisms and Treatment Considerations. International Journal of Molecular Sciences. 2023;24. doi: 10.3390/ijms24119568

10. Arnold KA, Blair JE, Paul JD, Shah AP, Nathan S, Alenghat FJ. Monocyte and macrophage subtypes as paired cell biomarkers for coronary artery disease. Experimental Physiology. 2019;104:1343–1352. doi: 10.1113/ep087827

11. Hou P, Fang J, Liu Z, Shi Y, Agostini M, Bernassola F, Bove P, Candi E, Rovella V, Sica G, et al. Macrophage polarization and metabolism in atherosclerosis. Cell Death & Disease. 2023;14. doi: 10.1038/s41419-023-06206-z

12. Barrett TJ. Macrophages in Atherosclerosis Regression. *Arteriosclerosis*, Thrombosis, and Vascular Biology. 2020;40:20–33. doi: 10.1161/atvbaha.119.312802

13. Liang X, Wang L, Wang M, Liu Z, Liu X, Zhang B, Liu E, Li G. MicroRNA-124 inhibits macrophage cell apoptosis via targeting p38/MAPK signaling pathway in atherosclerosis development. Aging (Albany NY). 2020;12:13005–13022. doi: 10.18632/aging.103387

14. Ben-Aicha S, Anwar M, Vilahur G, Martino F, Kyriazis PG, de Winter N, Punjabi PP, Angelini GD, Sattler S, Emanueli C. Small Extracellular Vesicles in the Pericardium Modulate Macrophage Immunophenotype in Coronary Artery Disease. JACC: Basic to Translational Science. 2024;9:1057–1072. doi: 10.1016/j.jacbts.2024.05.003

15. Zhang X, Rotllan N, Canfrán-Duque A, Sun J, Toczek J, Moshnikova A, Malik S, Price NL, Araldi E, Zhong W, et al. Targeted Suppression of miRNA-33 Using pHLIP Improves Atherosclerosis Regression. Circulation Research. 2022;131:77–90. doi: 10.1161/circresaha.121.320296

16. Mitchell PS, Parkin RK, Kroh EM, Fritz BR, Wyman SK, Pogosova-Agadjanyan EL, Peterson A, Noteboom J, O’Briant KC, Allen A, et al. Circulating microRNAs as stable blood-based markers for cancer detection. Proc Natl Acad Sci U S A. 2008;105:10513–10518. doi: 10.1073/pnas.0804549105

17. Weber JA, Baxter DH, Zhang S, Huang DY, Huang KH, Lee MJ, Galas DJ, Wang K. The microRNA spectrum in 12 body fluids. Clin Chem. 2010;56:1733–1741. doi: 10.1373/clinchem.2010.147405

18. Ikezaki H, Lim E, Cupples LA, Liu CT, Asztalos BF, Schaefer EJ. Small Dense Low-Density Lipoprotein Cholesterol Is the Most Atherogenic Lipoprotein Parameter in the Prospective Framingham Offspring Study. Journal of the American Heart Association. 2021;10. doi: 10.1161/jaha.120.019140

19. Sampson M, Wolska A, Warnick R, Lucero D, Remaley AT. A New Equation Based on the Standard Lipid Panel for Calculating Small Dense Low-Density Lipoprotein-Cholesterol and Its Use as a Risk-Enhancer Test. Clinical Chemistry. 2021;67:987–997. doi: 10.1093/clinchem/hvab048

20. Gai M-T, Yan S-Q, Wang M-y, Ruze A, Zhao L, Li Q-L, Zhao B-H, Deng A-X, Hu S, Gao X-M. Comparison of Gensini score and SYNTAX score for predicting in-stent restenosis in patients with coronary artery disease and drug-eluting stent implantation. Scientific Reports. 2025;15. doi: 10.1038/s41598-025-85191-z

21. Rampidis GP, Benetos G, Benz DC, Giannopoulos AA, Buechel RR. A guide for Gensini Score calculation. Atherosclerosis. 2019;287:181–183. doi: 10.1016/j.atherosclerosis.2019.05.012

22. Jiang J, Hiron TK, Agbaedeng TA, Malhotra Y, Drydale E, Bancroft J, Ng E, Reschen ME, Davison LJ, O’Callaghan CA. A Novel Macrophage Subpopulation Conveys Increased Genetic Risk of Coronary Artery Disease. Circulation Research. 2024;135:6–25. doi: 10.1161/circresaha.123.324172

23. Nagenborg J, Goossens P, Biessen EAL, Donners MMPC. Heterogeneity of atherosclerotic plaque macrophage origin, phenotype and functions: Implications for treatment. European Journal of Pharmacology. 2017;816:14–24. doi: 10.1016/j.ejphar.2017.10.005

24. Drechsler M, Duchene J, Soehnlein O. Chemokines Control Mobilization, Recruitment, and Fate of Monocytes in Atherosclerosis. *Arteriosclerosis*, Thrombosis, and Vascular Biology. 2015;35:1050–1055. doi: 10.1161/atvbaha.114.304649

25. Kang H, Li X, Xiong K, Song Z, Tian J, Wen Y, Sun A, Deng X, Garcia V. The Entry and Egress of Monocytes in Atherosclerosis: A Biochemical and Biomechanical Driven Process. Cardiovascular Therapeutics. 2021;2021:1–17. doi: 10.1155/2021/6642927

26. Zhou C, Gao Y, Ding P, Wu T, Ji G. The role of CXCL family members in different diseases. Cell Death Discovery. 2023;9. doi: 10.1038/s41420-023-01524-9

27. Li J, Dong Z, Pan Y, Wang L, Zhao W, Jian Z, M. A B. CPE Regulates Proliferation and Apoptosis of Primary Myocardial Cells Mediated by Ischemia and Hypoxia Injury. Journal of Healthcare Engineering. 2022;2022:1–9. doi: 10.1155/2022/3155171

28. Jia E-Z, Wang J, Yang Z-J, Zhu T-B, Wang L-S, Wang H, Li C-J, Chen BO, Cao K-J, Huang J, et al. Association of the mutation for the human carboxypeptidase E gene exon 4 with the severity of coronary artery atherosclerosis. Molecular Biology Reports. 2007;36:245–254. doi: 10.1007/s11033-007-9173-4

29. Wang J, Zhang Y, Yang Z-j, Zhu T-b, Wang L-s, Chen B, Cao K-j, Huang J, Ma W-z, Jia E-z. Association of human carboxypeptidase E exon5 gene polymorphisms with angiographical characteristics of coronary atherosclerosis in a Chinese population. Acta Pharmacologica Sinica. 2008;29:736–744. doi: 10.1111/j.1745-7254.2008.00798.x

30. Zuo Z, Zhou Z, Chang Y, Liu Y, Shen Y, Li Q, Zhang L. Ribonucleotide reductase M2 (RRM2): Regulation, function and targeting strategy in human cancer. Genes & Diseases. 2024;11:218–233. doi: 10.1016/j.gendis.2022.11.022

31. Tang B, Xu W, Wang Y, Zhu J, Wang H, Tu J, Weng Q, Kong C, Yang Y, Qiu R, et al. Identification of critical ferroptosis regulators in lung adenocarcinoma that RRM2 facilitates tumor immune infiltration by inhibiting ferroptotic death. Clinical Immunology. 2021;232. doi: 10.1016/j.clim.2021.108872

32. Lee SK, Hwang Y, Han JH, Haam S, Lee HW, Koh YW. Characteristics of the immune microenvironment associated with RRM2 expression and its application to PD-L1/PD-1 inhibitors in lung adenocarcinoma. Am J Cancer Res. 2023;13:5443–5454.

33. Maxwell KN, Breslow JL. Adenoviral-mediated expression of Pcsk9 in mice results in a low-density lipoprotein receptor knockout phenotype. Proc Natl Acad Sci U S A. 2004;101:7100–7105. doi: 10.1073/pnas.0402133101

34. Ellison M, Mittal M, Chaudhuri M, Chaudhuri G, Misra S. The role of the redox/miR-6855-3p/PRDX5A axis in reversing SLUG-mediated BRCA2 silencing in breast cancer cells. Cell Communication and Signaling. 2020;18. doi: 10.1186/s12964-019-0493-5

35. Abbas K, Breton J, Picot CR, Quesniaux V, Bouton C, Drapier JC. Signaling events leading to peroxiredoxin 5 up-regulation in immunostimulated macrophages. Free Radic Biol Med. 2009;47:794–802. doi: 10.1016/j.freeradbiomed.2009.06.018

36. Turchinovich A, Weiz L, Langheinz A, Burwinkel B. Characterization of extracellular circulating microRNA. Nucleic Acids Res. 2011;39:7223–7233. doi: 10.1093/nar/gkr254

37. Makarova JA, Shkurnikov MU, Wicklein D, Lange T, Samatov TR, Turchinovich AA, Tonevitsky AG. Intracellular and extracellular microRNA: An update on localization and biological role. Prog Histochem Cytochem. 2016;51:33–49. doi: 10.1016/j.proghi.2016.06.001

38. Sohel MH. Extracellular/Circulating MicroRNAs: Release Mechanisms, Functions and Challenges. Achievements in the Life Sciences. 2016;10:175–186. doi: 10.1016/j.als.2016.11.007

39. Swirski FK, Robbins CS, Nahrendorf M. Development and Function of Arterial and Cardiac Macrophages. Trends Immunol. 2016;37:32–40. doi: 10.1016/j.it.2015.11.004

40. Gupta RM, Lee-Kim VS, Libby P. The March of Monocytes in Atherosclerosis. Circulation Research. 2020;126:1324–1326. doi: 10.1161/circresaha.120.316981

41. Karhausen J, Bär F, Föh B, Pagel R, Schröder T, Schlichting H, Hirose M, Lemcke S, Klinger A, König P, et al. Carboxypeptidase E Modulates Intestinal Immune Homeostasis and Protects against Experimental Colitis in Mice. PLoS ONE. 2014;9. doi: 10.1371/journal.pone.0102347

42. Vishlaghi N, Guo L, Griswold-Wheeler D, Sun Y, Booker C, Crossley JL, Bancroft AC, Juan C, Korlakunta S, Ramesh S, et al. Vegfc-expressing cells form heterotopic bone after musculoskeletal injury. Cell Rep. 2024;43:114049. doi: 10.1016/j.celrep.2024.114049

43. Wang Q, Ismahil MA, Zhu Y, Rokosh G, Hamid T, Zhou G, Pogwizd SM, Prabhu SD. CD206(+)IL-4Ralpha(+) Macrophages Are Drivers of Adverse Cardiac Remodeling in Ischemic Cardiomyopathy. Circulation. 2025;152:257–273. doi: 10.1161/CIRCULATIONAHA.124.072411

44. Kim K, Shim D, Lee JS, Zaitsev K, Williams JW, Kim KW, Jang MY, Seok Jang H, Yun TJ, Lee SH, et al. Transcriptome Analysis Reveals Nonfoamy Rather Than Foamy Plaque Macrophages Are Proinflammatory in Atherosclerotic Murine Models. Circ Res. 2018;123:1127–1142. doi: 10.1161/CIRCRESAHA.118.312804

45. Liu Y, Zhong C, Chen S, Xue Y, Wei Z, Dong L, Kang L. Circulating exosomal mir-16-2-3p is associated with coronary microvascular dysfunction in diabetes through regulating the fatty acid degradation of endothelial cells. Cardiovasc Diabetol. 2024;23:60. doi: 10.1186/s12933-024-02142-0

46. Tian Z, Ning H, Wang X, Wang Y, Han T, Sun C. Endothelial Autophagy Promotes Atheroprotective Communication Between Endothelial and Smooth Muscle Cells via Exosome-Mediated Delivery of miR-204-5p. Arterioscler Thromb Vasc Biol. 2024;44:1813–1832. doi: 10.1161/ATVBAHA.123.319993

47. Hao X, Zhao B, Towers M, Liao L, Monteiro EL, Xu X, Freeman C, Peng H, Tang HY, Havas A, et al. TXNRD1 drives the innate immune response in senescent cells with implications for age-associated inflammation. Nat Aging. 2024;4:185–197. doi: 10.1038/s43587-023-00564-1

48. Yu J, Long B, Li Z, Tian X, Li D, Long J, Wang Y, Chen Y, Zhang F, Liu H, et al. Central memory CD4+ T cells play a protective role against immune checkpoint inhibitor-associated myocarditis. Cardiovasc Res. 2024;120:1442–1455. doi: 10.1093/cvr/cvae133

49. Wu J, Zhang Y, Tang H, Ye BC. MicroRNA-144-3p Inhibits Host Lipid Catabolism and Autophagy by Targeting PPARalpha and ABCA1 During Mycobacterium Tuberculosis Infection. ACS Infect Dis. 2024;10:1654–1663. doi: 10.1021/acsinfecdis.3c00731

